# Automated Sleep Staging Performance Across Two Experimental Sleep Manipulation Protocols in Adolescents

**DOI:** 10.64898/2026.10.24.26366382

**Authors:** João Paulo Lima Santos, London Evans, Rebecca L. Griffith, Shaun M. Purcell, Maria Jalbrzikowski, Peter Franzen, Adriane M. Soehner

## Abstract

Automated sleep staging algorithms improve the scalability of polysomnography analysis in adolescent research but are primarily validated in undisturbed, single-night sleep recordings. It remains unclear whether these algorithms maintain performance when sleep architecture is experimentally manipulated. This study evaluated the validity of two popular automated sleep staging approaches across two distinct sleep manipulation paradigms in adolescents: a sleep restriction protocol (N=43, mean age [SD]=13.37[1.06] years; four nights including two sleep extension [10hr] and two restriction [4hr] nights) and a slow-wave sleep deprivation (SWSD) protocol (N=18, mean age [SD]=17.17[1.79] years; baseline, SWSD, and recovery nights). Automated sleep staging was performed using the POPulation-level Sleep Stager (POPS), the Yet Another Spindle Algorithm (YASA), and combined POPS+YASA. Manual polysomnography scoring by experienced sleep technicians served as the reference standard. Agreement was assessed using Cohen’s κ, and stage-specific performance was evaluated using F1 scores and confusion matrices. Mixed-effects models tested differences in overall and stage-specific agreement across nights. All staging approaches demonstrated moderate-to-substantial agreement with manual scoring across manipulated sleep conditions (κ range=0.53-0.76). In the sleep restriction study, agreement was lower in restricted versus extended sleep (P<0.05), and agreement declined across restriction nights, with poorer Wake, N1, and N2 classification, while N3 remained preserved. In the SWSD study, overall agreement was lower on the SWSD night, with poorer N3 classification while other stages were preserved. Automated sleep staging performance may be less consistent in adolescents when sleep is curtailed or fragmented; future studies should develop and validate these algorithms under multiple sleep conditions.

## 1. INTRODUCTION

Sleep staging is a foundational component of human sleep research, providing the basis for quantifying sleep architecture and linking sleep physiology to cognitive, emotional, and health outcomes(Altevogt & Colten, 2006; Panel et al., 2015; Watson et al., 2015). According to the American Academy of Sleep Medicine (AASM) classification standards, manual scoring of polysomnography (PSG) recordings into Wake, N1, N2, N3, and REM sleep is the gold standard for sleep staging(Berry, 2014; Rechtschaffen, 1968). However, this process is labor-intensive, requiring trained scorers and several hours to score a single night of recording, limiting scalability in large studies. Automated sleep staging algorithms have been developed to address many of these limitations, with most algorithms trained and validated on undisturbed, single-night sleep recordings(Hanif et al., 2025; Vallat & Walker, 2021). Yet, it remains largely unknown whether these algorithms maintain comparable performance in experimental paradigms that deliberately manipulate sleep architecture. This limitation is important because inaccurate staging could obscure the effects of experimental manipulations on sleep architecture.

Experimental sleep manipulation paradigms are used to probe sleep regulation and its functional consequences by altering aspects of sleep architecture(Lima Santos et al., 2025; McMakin et al., 2016; Reddy et al., 2017). These paradigms include changes in total sleep opportunity, such as sleep restriction and sleep extension, as well as more targeted manipulations of sleep stages or sleep continuity and alterations in sleep timing or circadian alignment(Carrington & Trinder, 2008; Lima Santos et al., 2025; Palatine et al., 2024; Zhou et al., 2011). Such manipulations alter sleep architecture in different ways but ultimately produce substantial changes in the temporal organization and proportional composition of sleep stages across the sleep opportunity. These altered sleep patterns may be underrepresented in datasets used to train and validate automated staging algorithms and may limit their generalizability when the typical temporal organization and relative distribution of sleep stages are deliberately altered. This issue is also relevant to naturalistic sleep research using home recordings, where sleep patterns may vary more substantially than under controlled laboratory conditions, underscoring the importance of evaluating automated staging algorithms across under more variable sleep conditions. Experimental sleep manipulations provide a controlled framework for examining whether automated staging performance is sensitive to such changes in sleep architecture.

However, to the best of our knowledge, only one study has examined whether automated sleep staging algorithms remain robust under experimental sleep manipulation. A recent study in healthy adults(Chen et al., 2026) reported poorer performance of a commonly used algorithm (Yet Another Spindle Algorithm; YASA)(Vallat & Walker, 2021) during sleep restriction nights. Participants underwent laboratory-based PSG across adaptation, normal-sleep, and sleep-restriction conditions(Chen et al., 2026). Following an adaptation night, participants completed three consecutive nights of normal sleep (22:00-7:00) approximately one week later, followed by three consecutive nights of sleep restriction (1:30-7:00) after a four-day washout period(Chen et al., 2026). The order of the normal-sleep and sleep-restriction conditions was counterbalanced(Chen et al., 2026). The authors observed reduced agreement between manual and automated scoring for Wake, N1, and REM sleep in the sleep restriction condition, while agreement for N2 and N3 remained relatively preserved(Chen et al., 2026). However, because only the YASA algorithm was evaluated, we do not yet understand whether poorer agreement represents a general limitation of automated sleep staging under experimentally manipulated sleep, or an effect specific to this algorithm. Establishing whether these effects generalize across algorithms and sleep-manipulation paradigms is important because, if algorithm performance varies systematically under different manipulations, apparent changes in sleep-stage distribution could partly reflect measurement error rather than true physiological changes. This gap is particularly relevant for adolescent populations, for which fewer validation studies of automated sleep staging algorithms are available. Evaluating algorithms across distinct manipulation paradigms in adolescents is therefore necessary to determine whether reduced agreement is algorithm-specific or reflects a broader limitation of automated sleep staging.

This study aimed to evaluate the performance of two popular automated sleep staging algorithms in the context of experimental sleep manipulations relative manual PSG scoring by experienced technicians serving as the reference standard. Specifically, we examine the performance of the POPulation-level Sleep Stager (POPS)(Hanif et al., 2025; Purcell, 2026) and the YASA(Vallat & Walker, 2021). To assess algorithm performance across distinct forms of sleep manipulation, we analyze data from two experimental adolescent studies: a sleep restriction protocol that manipulates total sleep opportunity and a slow-wave sleep deprivation (SWSD) protocol that selectively targets slow-wave sleep while preserving overall sleep opportunity. We hypothesize that both algorithms will demonstrate poorer agreement with manual scoring during manipulated sleep conditions compared with non-manipulated nights. We further hypothesize that sleep restriction will affect agreement across multiple sleep stages, whereas SWSD will produce more selective reductions in staging performance, primarily involving SWS (N3).

## 2. METHODS

### 2.1. Participants

The present study was a secondary data analysis of overnight PSG data from two experimental studies that manipulated sleep opportunity and sleep architecture. Parental informed consent and adolescent assent were obtained. Both studies were approved by the Institutional Review Board at the University of Pittsburgh.

#### Study 1 – Sleep restriction vs extension

The first dataset was a sleep restriction vs. sleep extension protocol study designed to investigate the effects of sleep loss on neurobehavioral reward and emotion regulation in typically developing youth ages 11-15 years-old(McMakin et al., 2016). Of the 57 participants enrolled, 43 had complete PSG recordings available for all four nights. Participants were required to have no psychiatric disorders.

#### Study 2 – Slow wave sleep deprivation (SWSD)

The second dataset was a pilot study of the antidepressant and neurobehavioral effects of SWSD in adolescents 12-19 years-old with depressive symptoms(Lima Santos et al., 2025; Palatine et al., 2024). A total of 18 participants were enrolled, all of whom had complete PSG recordings available for all three nights. Participants were required to have at least mild depressive symptoms in the past 2 weeks (Patient Health Questionnaire-9 ≥ 5).

Common exclusion criteria for both studies included history of psychosis or bipolar disorder, substance/alcohol dependence or abuse, sleep disorders, neurological or major systemic medical conditions, history of head trauma, and use of medications that could affect the central nervous system. Study-specific exclusion criteria are detailed in the Supplemental Materials. Sample characteristics are shown in Table 1.

**Table 1.** Demographic characteristics of the studies.

| Variable | Sleep restriction study<br>(N=43) <sup>a</sup> | SWSD study<br>(N=18) |
| --- | --- | --- |
| Age (years), mean [SD] | 13.37 [1.06] | 17.17 [1.79] |
| Sex at birth |  |  |
| Male, N(%) | 19 (44%) | 4 (22%) |
| Female, N(%) | 24 (56%) | 14 (78%) |
| Ethnicity |  |  |
| Hispanic, N(%) | 2 (5%) | 1 (6%) |
| Non-hispanic, N(%) | 40 (93%) | 17 (94%) |
| Race |  |  |
| Asian, N(%) | 1 (2%) | 5 (28%) |
| Black, N(%) | 5 (12%) | 3 (17%) |
| Multiple, N(%) | 1 (2%) | 1 (6%) |
| White, N(%) | 33 (77%) | 9 (50%) |
| Depression Severity (MFQ), mean [SD] | 2.23 [1.6] <sup>b</sup> | 25.3 (13.9) |
| History of depression, N(%) | 0 (0%) | 18 (100%) |

<sup>a</sup> One participant did not report ethnicity, and three participants did not report race. Abbreviations: MFQ – Mood and Feelings Questionnaire
<sup>b</sup> The MFQ was only partially collected; data were available for only 21 participants.

### 2.2. PSG Data Collection and Processing

#### 2.2.1. PSG data collection

In both studies, PSG recordings included bilateral frontal, central, parietal, and occipital EEG leads referenced to the contralateral mastoids(Brindle et al., 2018). Study 1 collected PSG data using the Vitaport and Compumedics Grael systems, whereas Study 2 used the Compumedics Grael system.

#### 2.2.2. Manual sleep staging

PSG recordings were manually scored in 30-second (s) epochs according to AASM(Iber, 2007) by trained scorers at the University of Pittsburgh Sleep Laboratory. Inter-rater reliability across scorers in the Sleep Laboratory has averaged above 92% over the past 12 years, above the laboratory’s 90% benchmark. The sleep-stage label assigned to each 30-s epoch was extracted for each recording for use in the main analyses. In addition, the total duration of each sleep stage (N1, N2, N3, and REM) was extracted for each recording for use in exploratory analyses.

#### 2.2.3. Automated sleep staging algorithms

Automated sleep staging was performed using two open-access algorithms: YASA and the POPS. Both use time- and frequency-domain features for automated classification and were trained and evaluated using the National Sleep Research Resource (NSRR) dataset. This shared training and testing dataset provided a common basis for comparing their performance under experimentally manipulated sleep conditions and assessing whether observed changes were consistent across the two approaches.

YASA is an open-source automated sleep staging pipeline trained on more than 30,000 hours of PSG recordings. YASA uses a single central EEG channel to extract time- and frequency-domain features from consecutive 30-s epochs and applies a machine-learning classifier to assign each epoch to an AASM sleep stage (Wake, N1, N2, N3, or REM) (Vallat & Walker, 2021).

POPS is Luna’s automated sleep staging pipeline (the open-source package used for preprocessing; see Section 2.4)(Purcell, 2026). For the single-channel model, POPS derives epoch-level time- and frequency-domain features and applies a gradient-boosted decision-tree classifier (LightGBM) to generate posterior probabilities for the five standard AASM sleep stages (Wake, N1, N2, N3, and REM) for each 30-s epoch(Purcell, 2026).

#### 2.2.4. Automated sleep staging preprocessing

The EEG recordings for the SWSD study included a 0.3 Hz low-frequency filter, a 60 Hz notch filter, and a 35 Hz high-frequency filter. The EEG recordings for the Sleep Restriction study were provided without a band-pass filter but had the same 60 Hz notch filter; therefore, a 0.3–35 Hz band-pass filter was applied during preprocessing to harmonize filtering across datasets. Data were processed using the open-source package Luna(Purcell, 2026). Original EEG recordings were resampled at 128 Hz. Analyses focused on the C3-M2 and C4-M1 derivations, which were available across both datasets. Artifact detection and removal were performed within N2, N3, and REM epochs using predefined criteria(Hjorth, 1970): epochs were identified as containing artifacts if they met any of the following criteria: (a) delta power greater than 2.5 times the local average or beta power greater than 2.0 times the local average, calculated using Welch’s method; (b) maximum amplitudes exceeding 250 μV for more than 5% of the epochs; (c) flat or clipped signals for more than 5% of the epochs; or (d) signals exceeding 3.5 standard deviations from the mean for any of the three Hjorth parameters (activity, mobility, or complexity)(Hjorth, 1970). Outlier removal based on Hjorth parameters was performed twice for each recording, following prior work(Kozhemiako et al., 2024). Channels were excluded if more than 50% of epochs contained identified artifacts. Records with less than 180 min of total sleep time (TST) were also excluded, as these recordings may reflect atypical sleep conditions unrelated to the experimental sleep manipulation.

Sleep staging for each recording was performed independently using both POPS and YASA. For each algorithm, a best-channel staging output was generated by selecting, for each epoch, the prediction from the EEG derivation (C3-M2 or C4-M1) with the highest model confidence score (posterior probability). This approach leveraged the channel with the greatest model-estimated confidence on a scale of 0-1 at each time point. A POPS+YASA combined staging output was subsequently generated by selecting, for each epoch, the prediction with the highest confidence score across the best-channel outputs from both algorithms. This procedure yielded three staging outputs for each recording: POPS, YASA, and Combined.

For each automated algorithm, the sleep-stage label assigned to each 30-s epoch was extracted for each recording for use in the main analyses. In addition, the total duration of each sleep stage (N1, N2, N3, and REM) was extracted for each recording for use in exploratory analyses.

### 2.3. Sleep manipulation procedures

#### 2.3.1. Sleep restriction vs extension study

Participants completed two 48-hour in-laboratory visits during the summer months, with visits separated by approximately one week. The order of sleep restriction versus extension was counterbalanced across participants. During the sleep restriction condition, participants were limited to 4 hours of sleep opportunity per night (01:00-05:00; Restriction nights) for two consecutive nights. During the sleep extension condition, participants received 10 hours of sleep opportunity per night (22:00-08:00; Extension nights) for two consecutive nights. Sleep opportunities for sleep restriction and extension were centered on the same sleep midpoint. Participants were continuously monitored during laboratory visits to ensure wakefulness outside of scheduled sleep periods. Sleep was recorded with PSG overnight.

#### 2.3.2. SWSD study

Participants were instructed to maintain a stable sleep schedule at their habitual sleep times with an 8-hour sleep opportunity during the week preceding the sleep laboratory visit, which was verified with sleep diary and actigraphy. Participants completed a 3-night in-laboratory polysomnographic protocol, consisting of one Baseline night (Night 1), one selective SWSD night (Night 2), and one Recovery night (Night 3). Sleep and wake times during laboratory visits were scheduled according to participants’ habitual sleep schedules. During baseline and recovery nights, participants slept undisturbed. During the manipulation night, acoustic stimulation was delivered in real time based on sleep EEG monitoring to selectively disrupt slow-wave sleep(Lima Santos et al., 2025; Palatine et al., 2024). Sleep was recorded with PSG overnight. While all participants received SWSD on night 2, they were informed that they might either be allocated to a Sham condition or receive SWSD on night 2, to reduce the effects of expectancies on primary mood and behavioral outcomes.

### 2.4. Statistical approach

Automated sleep staging performance was evaluated separately for each study (Sleep restriction or SWSD) and algorithm (POPS, YASA, or Combined), using manual sleep staging as the reference. Analyses were conducted at both the study level and participant level to characterize overall agreement and examine changes in performance across nights.

#### 2.4.1. Study-level agreement between manual vs automated sleep staging

Agreement between manual and automated sleep staging was quantified using Cohen’s κ(McHugh, 2012), a chance-corrected measure of agreement for categorical data. κ ranges from - 1 to 1, where 0 indicates agreement equivalent to chance, negative values indicate agreement worse than chance, and positive values indicate increasing levels of agreement, with 1 indicating perfect agreement. For descriptive purposes, positive κ values were interpreted using conventional benchmarks: <0.20 (slight), 0.21-0.40 (fair), 0.41-0.60 (moderate), 0.61-0.80 (substantial), and >0.81 (almost perfect agreement)(Landis & Koch, 1977).

Stage-specific classification performance was assessed using F1 scores for Wake, N1, N2, N3, and REM sleep. The F1 score is the harmonic mean of precision and recall, providing a robust measure of performance in the presence of class imbalance(Powers, 2020). F1 scores range from 0 to 1, with higher values indicating better correspondence between automated predictions and manual sleep stage labels. Confusion matrices were also generated to characterize common patterns of misclassification. These metrics were calculated separately for each algorithm and study night.

#### 2.4.2. Participant-level agreement between manual vs automated sleep staging

To evaluate whether sleep staging performance differed across nights, we used mixed-effect models. Cohen’s κ and stage-specific F1 scores were treated as the dependent variables, with study night as the independent variable (fixed effect). Separate mixed-effects models were fitted for Cohen’s κ and the F1 score of each sleep stage. Participant was included as a random effect. Pairwise contrasts were then used to compare the performance between each pair of study nights within each study. For the Sleep Restriction study, all pairwise comparisons among Extension night 1, Extension night 2, Restriction night 1, and Restriction night 2 were performed, resulting in six comparisons. For the SWSD study, all pairwise comparisons among Baseline night, SWSD night, and Recovery night were performed, resulting in three comparisons. False Discovery Rate (FDR) correction was used to correct for multiple comparison(Benjamini & Hochberg, 1995).

#### 2.4.3. Exploratory analyses

##### Additional performance metrics

Sensitivity and specificity were calculated for Wake, N1, N2, N3, and REM to further characterize stage-specific classification performance relative to manual scoring across nights.

##### Correlation between manual and automated sleep stages duration

Pearson correlations were calculated between automated and manual sleep-stage duration estimates for each study, night, sleep stage, and algorithm. Steiger’s test was used to compare correlations between nights, with FDR correction for multiple comparisons.

## 3. RESULTS

### 3.1. Overall characteristics

All participants’ sleep recordings passed quality control and were therefore included in the models. Demographic characteristics are described by study in Table 1.

Supplemental Table 1 describes sleep stages by night for each study. In the sleep restriction study, N3% increased progressively across Restriction nights (53.65% and 62.02%), accompanied by reductions in N2 (23.92% and 19.47%) and REM sleep (10.61% and 13.27%). N1 sleep decreased across Restriction nights, while Wake showed a reduction primarily during the second Restriction night compared with Extension nights.

In the SWSD study, manual scoring demonstrated a selective reduction in N3% during the SWSD night (8.44%) compared with the Baseline (22.20%) and Recovery (24.04%) nights. In contrast, N2 increased during SWSD (54.04%) compared with the Baseline (42.27%) and Recovery (42.03%) nights. Wake, N1, and REM sleep showed minimal changes across conditions, with the Baseline and Recovery nights showing comparable sleep stage distributions (Supplemental Table 1).

### 3.2. Sleep restriction study (Study 1)

#### 3.2.1. Study-level agreement

Cohen’s κ indicated **substantial** agreement between manual and automated staging across all algorithms on both Extension nights (Extension night 1: POPS κ=0.70; YASA κ=0.68, Combined κ=0.70; Extension night 2: POPS κ=0.69; YASA κ=0.67, Combined κ=0.69; Table 2). Stage-wise F1 scores were highest for REM, N3, Wake, and N2, reflecting strong correspondence across algorithms. N1 showed the lowest F1 scores in all algorithms on both Extension nights, with confusion matrices indicating that N1 was primarily misclassified as Wake (Table 2; Supplementary Table 2).

**Table 2.** Inter-rater agreement and stage specific performance.

| **Sleep restriction study** |  |  |  |  |  |  |  |
| --- | --- | --- | --- | --- | --- | --- | --- |
| Night | Algorithm | Cohen's κ<sup>1</sup> | F1 scores for each sleep stage | | | | |
|  |  |  | Wake | N1 | N2 | N3 | REM |
| Extension night 1 | POPS | **0.70** | 0.82 | 0.29 | 0.75 | 0.80 | 0.85 |
|  | YASA | **0.68** | 0.78 | 0.30 | 0.75 | 0.78 | 0.84 |
|  | Combined | **0.70** | 0.79 | 0.31 | 0.76 | 0.79 | 0.86 |
| Extension night 2 | POPS | **0.69** | 0.77 | 0.26 | 0.75 | 0.80 | 0.85 |
|  | YASA | **0.67** | 0.74 | 0.26 | 0.75 | 0.78 | 0.83 |
|  | Combined | **0.69** | 0.75 | 0.28 | 0.76 | 0.79 | 0.85 |
| Restriction night 1 | POPS | **0.61** | 0.79 | 0.21 | 0.60 | 0.80 | 0.79 |
|  | YASA | **0.61** | 0.73 | 0.17 | 0.66 | 0.79 | 0.82 |
|  | Combined | **0.62** | 0.76 | 0.18 | 0.65 | 0.79 | 0.84 |
| Restriction night 2 | POPS | **0.54** | 0.64 | 0.17 | 0.50 | 0.82 | 0.76 |
|  | YASA | **0.53** | 0.54 | 0.09 | 0.55 | 0.81 | 0.74 |
|  | Combined | **0.56** | 0.57 | 0.13 | 0.56 | 0.82 | 0.78 |
| **SWSD study** |  |  |  |  |  |  |  |
| Night | Algorithm | Cohen's $\kappa$ | F1 scores for each sleep stage | | | | |
|  |  |  | Wake | N1 | N2 | N3 | REM |
| Baseline night | POPS | **0.73** | 0.68 | 0.20 | 0.85 | 0.88 | 0.82 |
|  | YASA | **0.76** | 0.79 | 0.26 | 0.85 | 0.87 | 0.85 |
|  | Combined | **0.76** | 0.73 | 0.25 | 0.86 | 0.89 | 0.85 |
| SWSD night | POPS | **0.67** | 0.66 | 0.25 | 0.87 | 0.58 | 0.84 |
|  | YASA | **0.66** | 0.71 | 0.31 | 0.83 | 0.55 | 0.88 |
|  | Combined | **0.68** | 0.69 | 0.26 | 0.85 | 0.58 | 0.88 |
| Recovery night | POPS | **0.72** | 0.62 | 0.22 | 0.86 | 0.91 | 0.76 |
|  | YASA | **0.76** | 0.74 | 0.30 | 0.86 | 0.90 | 0.82 |
|  | Combined | **0.76** | 0.68 | 0.22 | 0.87 | 0.91 | 0.81 |

Abbreviations: POPS – POPulation level Sleep Stager; YASA -Yet Another Spindle Algorithm; SWSD - Slow-Wave Sleep Deprivation.
<sup>1</sup> Cohen's $\kappa$ values benchmarks: <0.20 (slight), 0.21-0.40 (fair), 0.41-0.60 (moderate; in **red**), 0.61-0.80 (substantial; in **blue**), and >0.81 (almost perfect agreement).

On Restriction night 1, Cohen’s κ indicated **substantial** agreement between manual and automated staging across all algorithms (POPS κ=0.61; YASA κ=0.61, Combined κ=0.62; Table 2). Stage-wise F1 scores were highest for REM, N3, and Wake, while N2 showed intermediate correspondence and N1 remained the lowest. Confusion matrices showed that N1 was primarily misclassified as Wake, whereas N2 misclassifications occurred across multiple stages (Table 2; Supplementary Table 3).

On Restriction night 2, agreement further declined to **moderate** levels (POPS κ=0.54; YASA κ=0.53, Combined κ=0.56; Table 2). Stage-wise F1 scores were highest for REM and N3, while Wake and N2 showed intermediate correspondence and N1 remained the lowest. N1 was primarily misclassified as Wake, whereas N2 errors remained distributed across multiple stages (Table 2; Supplementary Table 3).

#### 3.2.2. Per-subject contrasts across nights

Across Extension nights 1 and 2, mixed-effects models revealed no significant difference between nights (Figure 1A) and no stage-specific effects (Figure 2A; Supplementary Tables 4-5).

**Figure 1.**
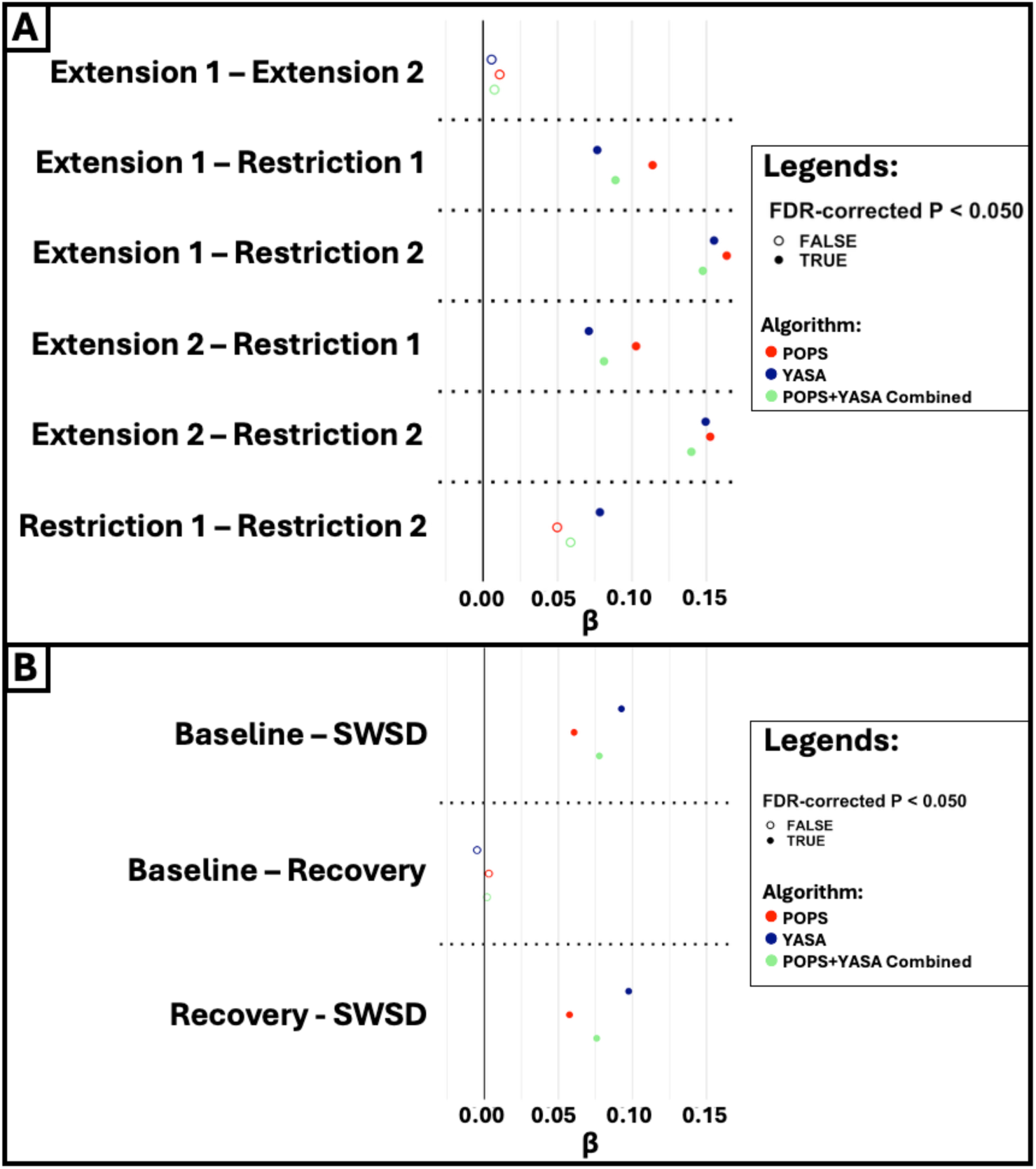
Differences in sleep staging agreement across nights and algorithms in the Sleep restriction and SWSD studies. Panel A shows results from the Sleep restriction study, and Panel B shows results from the Slow-Wave Sleep Deprivation (SWSD) study. The x-axis displays regression coefficients (β) from mixed-effects models quantifying differences in Cohen’s κ across nights. The y-axis lists pairwise night comparisons within each study. Colors indicate sleep staging algorithm (POPS: red; YASA: blue; Combined: green). Filled circles denote statistically significant effects after false discovery rate (FDR) correction (P<0.05), whereas open circles indicate non-significant effects. Horizontal dotted lines separate individual contrast levels for visual clarity. Positive β values indicate higher agreement (Cohen’s κ) in the first condition of each contrast relative to the second, whereas negative β values indicate lower agreement.

**Figure 2.**
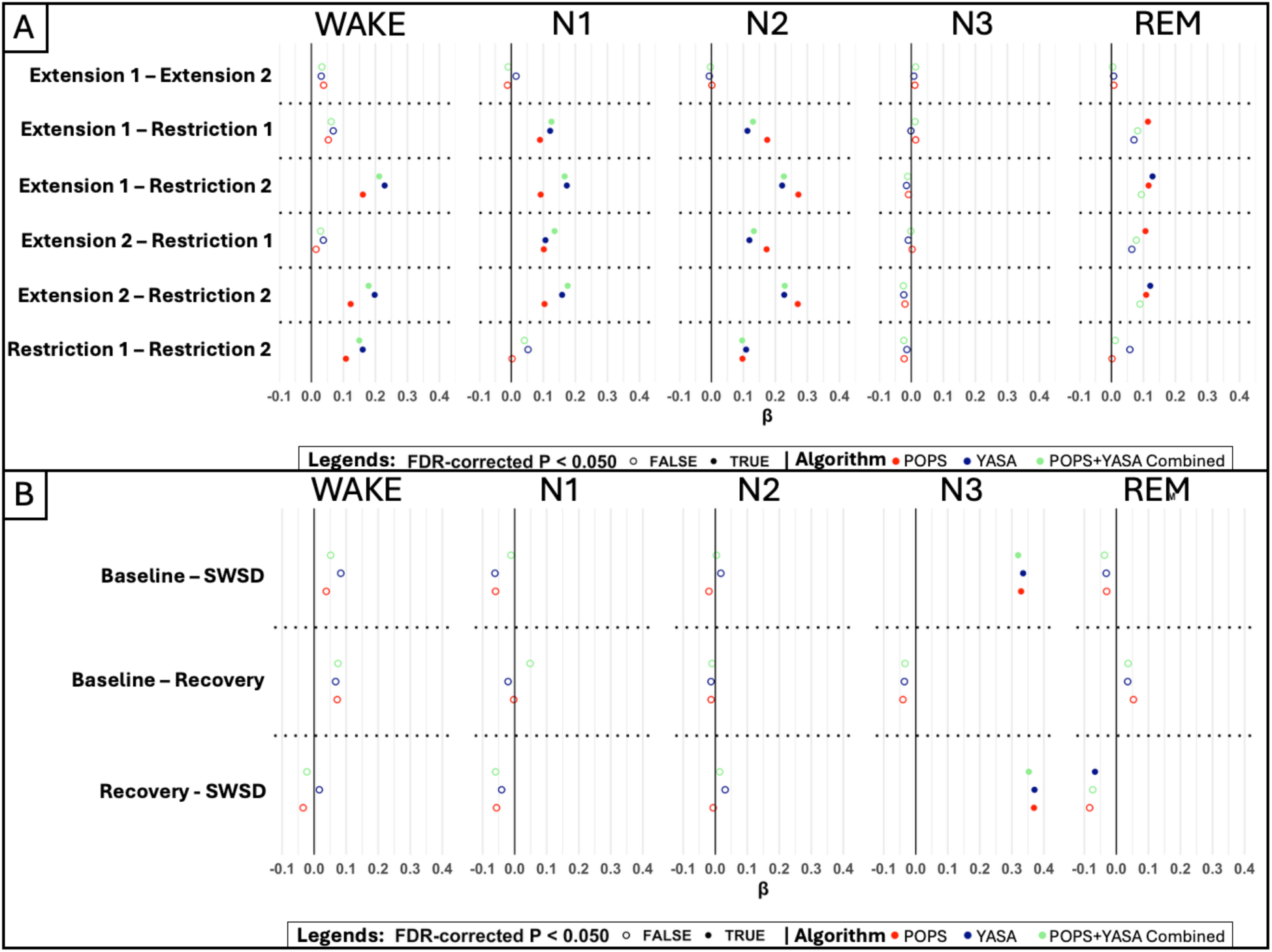
Stage-specific differences in automated sleep staging performance across nights and algorithms. Panel A shows results from the Sleep restriction study, and Panel B shows results from the Slow-Wave Sleep Deprivation (SWSD) study. The x-axis displays regression coefficients (β) from mixed-effects models quantifying differences in stage-specific F1 scores across experimental nights. Positive β values indicate higher F1 scores (i.e., better agreement) in the first condition of each contrast relative to the second condition, whereas negative β values indicate lower F1 scores (i.e., poorer agreement). The y-axis indicates pairwise night contrasts within each study. Each facet corresponds to a sleep stage (Wake, N1, N2, N3, and REM). Colors indicate automated sleep staging algorithm (POPS: red; YASA: blue; Combined: green). Filled circles denote statistically significant effects after false discovery rate (FDR) correction (P < 0.05), whereas open circles indicate non-significant effects. Horizontal dotted lines separate contrast levels for visual clarity.

In contrast, comparisons between Extension and Restriction nights revealed significantly lower κ during Restriction nights, indicating reduced manual vs automated staging agreement under sleep restriction (Figure 1A; Supplementary Table 4). Stage-specific analyses showed that N3 detection was unaffected across all algorithms, whereas N1 and N2 detection were consistently reduced. REM detection was affected in POPS and YASA, but not in the Combined algorithm. Wake detection remained stable on the first night of sleep restriction but was reduced on the second night (Figure 2A; Supplementary Table 5).

Finally, comparisons between Restriction night 1 and Restriction night 2 revealed a significant decline in agreement for YASA only, whereas POPS and Combined remained stable across restriction nights (Figure 1A; Supplementary Table 4). In addition, Wake and N2 detection showed a significant reduction in agreement from Restriction night 1 to Restriction night 2 across all algorithms, whereas N1, N3, and REM did not differ significantly between Restriction nights (Figure 2A; Supplementary Table 5).

### 3.3. SWSD study (Study 2)

#### 3.3.1. Study-level agreement

For the Baseline night, Cohen’s κ indicated **substantial** agreement between manual and automated staging across all algorithms (POPS κ=0.73; YASA κ=0.76, Combined κ=0.76; Table 2). Stage-wise F1 scores were highest for REM, N3, and N2, while Wake showed intermediate correspondence. N1 showed the lowest F1 scores in all algorithms, with confusion matrices indicating that N1 was primarily misclassified as Wake (Table 2; Supplementary Table 6).

For the SWSD night, Cohen’s κ indicated **substantial** agreement between manual and automated staging across all algorithms (POPS κ=0.67; YASA κ=0.66, Combined κ=0.68; Table 2). Stage-wise F1 scores were highest for REM, and N2, while Wake and N3 showed intermediate correspondence. N1 showed the lowest F1 scores in all algorithms. Confusion matrices indicated that N1 was primarily misclassified as Wake or N2 while N3 was primarily misclassified as N2 (Table 2; Supplementary Table 6).

For the Recovery night, Cohen’s κ indicated **substantial** agreement between manual and automated staging across all algorithms (POPS κ=0.72; YASA κ=0.76, Combined κ=0.76; Table 2). Stage-wise F1 scores were highest for REM, N3, and N2, while Wake showed intermediate correspondence. N1 showed the lowest F1 scores in all algorithms, with confusion matrices indicating that N1 was primarily misclassified as Wake (Table 2; Supplementary Table 6).

#### 3.3.2. Per-subject contrasts across nights

Mixed-effects analyses revealed that overall κ was significantly lower during SWSD night compared to both Baseline and Recovery nights (Figure 1B; Supplemental Table 7). Stage-specific analyses showed that Wake, N1, and N2 detection did not differ between SWSD night and either Baseline or Recovery nights (Figure 2B).

In contrast, N3 detection was significantly reduced during SWSD night relative to both Baseline and Recovery nights (Supplemental Table 8) for all three algorithms. For REM detection, no differences were observed between SWSD and Baseline nights or between Baseline and Recovery nights, except for the YASA algorithm, which showed slightly lower REM detection during the Recovery night compared to SWSD night (Supplemental Table 8).

Finally, no significant differences were found between Baseline and Recovery nights for overall κ or for any stage-specific metric (Figure 1B; Figure 2B; Supplemental Tables 7-8).

### 3.4. Exploratory analyses

#### Additional performance metrics

In the sleep restriction study, sensitivity generally decreased during restriction nights for N2 and REM, while N3 sensitivity remained relatively preserved. Specificity remained relatively stable across nights. In the SWSD study, N3 sensitivity decreased during the SWSD night, whereas N3 specificity remained relatively preserved. Sensitivity for the other sleep stages showed less consistent changes across nights, while specificity was generally stable (Supplemental Table 9).

#### Correlation between manual and automated sleep stages duration - Sleep Restriction study

Pearson correlations ranged from r = 0.154 to 0.827 across nights, sleep stages, and automated algorithms (Supplemental Figure 1). Correlations were positive and generally strongest for N2, N3, and REM, with lower associations for N1. Correlations were largely stable across nights; although several nominal differences were observed, none remained significant after FDR correction (Supplemental Table 10).

#### Correlation between manual and automated sleep stages duration - SWSD study

Pearson correlations ranged from r = −0.123 to 0.920 across nights, sleep stages, and automated algorithms (Supplemental Figure 2). Correlations were predominantly positive, with lower associations generally observed for N1. Correlations were largely stable across nights, except for N3, where correlations decreased during SWSD and increased during Recovery. The SWSD-to-Recovery change was significant for POPS (P = 0.002) and combined (P = 0.001) after FDR correction. No other comparisons remained significant after correction (Supplemental Table 11).

## 4. DISCUSSION

In this secondary data analysis, we evaluated the performance of three automated sleep staging approaches – POPS, YASA, and a Combined approach – relative to hand-scoring in two different sleep manipulation studies conducted with adolescents. Overall, all three automated approaches maintained substantial agreement with manual sleep staging across experimental conditions, supporting adequate performance when measuring manipulated sleep. However, automated staging results showed significantly lower agreement with manual staging on sleep restriction and SWSD nights, indicating that experimentally induced alterations in sleep architecture can impact staging accuracy. In addition, consecutive sleep restriction nights resulted in a progressive reduction in automated vs manual staging agreement, with lower correspondence observed during the second restriction night compared to the first. Importantly, automated sleep stage classification accuracy was not uniform across experimental paradigms.

In the sleep restriction protocol (Study 1), relative to manual scoring, all automated staging algorithms showed poorer classification accuracy for Wake, N1, N2, and REM during restriction nights, whereas N3 classification remained preserved. Notably, the stages showing reduced classification accuracy were also those that decreased in relative occurrence during sleep restriction, whereas N3 increased in occurrence while its classification accuracy remained preserved. The sensitivity and specificity analyses further indicate that these changes in classification performance were primarily reflected in sensitivity, while specificity remained relatively stable. Thus, changes in sleep-stage distribution alone cannot fully explain the degradation in automated staging performance; rather, the direction of change in stage occurrence may be relevant, with reductions in the representation of a stage potentially making it more difficult to detect. This interpretation is further supported by the SWSD protocol (Study 2), in which selective reduction of N3 was accompanied by impaired N3 classification, whereas classification of stages that were preserved remained similar across nights. Together, these findings suggest that automated staging approaches may be particularly sensitive to reductions in the representation of specific sleep stages, rather than to changes in sleep architecture alone. These findings may reflect the physiological and temporal information used by automated algorithms to distinguish sleep stages. For example, YASA(Vallat & Walker, 2021) incorporates spectral and time-domain EEG features as well as information from neighboring epochs, indicating that classification depends on both stage-specific signal characteristics and the temporal context in which an epoch occurs. Changes in these characteristics or in the temporal context of a stage may therefore affect its classification, particularly when that stage becomes less prevalent.

Our findings are consistent with previous work evaluating YASA during sleep restriction in adults, which reported reduced staging performance across multiple sleep stages(Chen et al., 2026). Our study extends these observations to adolescents and demonstrates that reduced performance under altered sleep architecture is not limited to a single algorithm, but is observed across multiple automated staging approaches and distinct sleep manipulation paradigms. Moreover, the progressive decline in agreement across restriction nights suggests that cumulative sleep manipulation may further challenge automated staging performance, highlighting the importance of validating these approaches under conditions of sustained or repeated sleep disruption. However, it is important to acknowledge that manual scoring may also be impacted under sleep manipulation protocols. Although no direct work has investigated whether sleep restriction or SWSD affects manual staging reliability, manual sleep-stage scoring is itself subject to inter-scorer variability(Lee et al., 2022), and previous work has demonstrated variability in more complex sleep recordings, including those characterized by sleep fragmentation(Choi et al., 2026). Thus, changes in manual scoring reliability cannot be excluded as a potential contributor to the reduced automated-versus-manual agreement observed under sleep manipulation.

Consistent with previous reports, N1 remained the least accurately classified stage across both studies, reflecting the known difficulty of reliably distinguishing this transitional stage using automated approaches(Cesari et al., 2021; Sun et al., 2017). Additionally, although YASA and POPS showed comparable overall performance, the Combined approach demonstrated better performance, suggesting that combining approaches with complementary strengths and sources of error may help offset algorithm-specific weaknesses and improve classification when sleep architecture is altered. Finally, correlations between sleep-stage duration from automated algorithms and manual scoring were less affected by sleep manipulation than epoch classifications, with significant changes limited to N3 duration in the SWSD study after FDR correction. This suggests that, for SWSD, changes in epoch classification were sufficient to alter the overall estimate of N3 duration. In contrast, no significant changes in correlations were observed in the Sleep Restriction study after FDR correction, despite changes in epoch classification performance. Thus, sleep-stage duration alone may not fully capture changes in automated staging performance, highlighting the importance of evaluating both sleep-stage duration and epoch-level classification, with larger studies needed to confirm these findings.

While this work provides important insights into the performance of automated sleep staging under experimentally manipulated sleep conditions, several limitations should be considered. First, the sample sizes were relatively modest, particularly for the SWSD protocol, which may limit the generalizability of our findings. Second, both studies focused on adolescents; therefore, the extent to which these findings generalize to other age groups remains to be determined. Third, the sleep manipulations examined in these studies represent only a subset of the ways in which sleep may be disrupted. In particular, these protocols did not address disruptions in sleep timing or more general sleep fragmentation, which may differentially affect automated staging performance. Fourth, the SWSD protocol included a single night of selective sleep manipulation, and future studies incorporating longer or repeated manipulation periods will be important to determine whether staging performance is further impacted by sustained alterations in sleep architecture. Finally, we evaluated a limited set of automated staging algorithms and did not assess all approaches that have been validated in pediatric populations. Other algorithms developed and evaluated using different training datasets may show different patterns of performance under sleep manipulation. Future work should therefore evaluate a broader range of automated staging approaches across diverse populations, including both clinical and nonclinical cohorts, to determine whether similar stage-specific vulnerabilities emerge under different physiological and pathological conditions.

In conclusion, automated sleep staging approaches demonstrated adequate performance in adolescents; however, experimentally induced alterations in sleep architecture revealed stage-specific limitations in automated classification. Across sleep restriction and SWSD paradigms, classification performance was particularly affected when specific sleep stages were reduced or underrepresented, as demonstrated by impaired N3 detection during SWSD despite preserved performance when N3 was increased during sleep restriction. These findings highlight the importance of evaluating automated sleep staging algorithms across different sleep contexts, as performance that varies with sleep architecture under controlled experimental conditions may have implications for their use in naturalistic settings, where sleep patterns and environmental conditions are less controlled. Future work should investigate probabilistic approaches that retain information about stage-level uncertainty and dynamics, as well as evaluate automated staging in large-scale datasets encompassing a broader range of sleep conditions.

## ETHICS APPROVAL STATEMENT

This study was approved by the Institutional Review Board at the University of Pittsburgh.

## PATIENT CONSENT STATEMENT

Parental informed consent and adolescent assent were obtained.

## Data Availability

Data may be made available from the corresponding author upon reasonable request.

