## Supplemental materials for "Automated Sleep Staging Performance Across Two Experimental Sleep Manipulation Protocols in Adolescents"

**TITLE**:


**AUTHORS AND AFFILIATIONS**:

João Paulo Lima Santos[1], MD; London Evans[1], BA; Rebecca L. Griffith[1], PhD; Shaun M. Purcell[2], PhD; Maria Jalbrzikowski[3,4], PhD; Peter Franzen[1], PhD; Adriane M. Soehner[1]*, PhD.

1 Department of Psychiatry, University of Pittsburgh School of Medicine, Pittsburgh, PA, USA.

2 Department of Psychiatry, Columbia University Irving Medical Center - New York State Psychiatric Institute, New York, NY, USA.

3 Department of Psychiatry and Behavioral Sciences, Boston Children's Hospital, Boston, MA, USA.

4 Department of Psychiatry, Harvard Medical School, Boston, MA, USA.

**CORRESPONDING AUTHOR**:

*** Corresponding author:**

Adriane Soehner, PhD

Department of Psychiatry

University of Pittsburgh

3811 O'Hara St, E1329

Pittsburgh, PA 15213

### Supplemental methods

**Participants.**

Sleep restriction study (Study 1) – Exclusion criteria: (1) current or past history of psychiatric disorder; substance abuse; (2) current or past history of sleep disorder; (3) neurological or severe systemic medical illnesses; (4) self-reported average sleep duration < 6 hours or > 11 hours across weekday and weekend nights, or normal bedtime later than 1:00 am; (5) history of head trauma with loss of consciousness; (6) use of psychotropic medications, or medications for medical illness that produce CNS effects (e.g. steroids); (7) contraindications to MR scanning; (8) contraindications for sleep restriction (past history of seizure; first-degree relative with bipolar disorder); (9) visual disturbance; (10) non-right handedness; (11) being unable to complete questionnaires written in English; (12) Attending summer school during the period of summer participation.

SWSD study (Study 2) – Exclusion criteria: (1) history of psychosis or bipolar disorder; (2) use of psychiatric medication other than Selective Serotonin Reuptake Inhibitors (SSRI); (3) substance or alcohol dependance in the past 3 months; (4) intellectual disabilities; (5) a positive urine drug screening (other than cannabis); (6) self-reported hearing impairments; (7) pregnancy (only females); (8) sleep disorders that would make be an overnight sleep study challenging (e.g. chronic nightmares, severe insomnia) or organic sleep disorders (e.g., REM behavior disorder, narcolepsy) as per a locally-developed clinical interview; (9) neurological disorders, history of head trauma, or major systemic medical conditions.

### Supplemental tables

**Supplemental Table 1. Sleep stage distribution (percentage) based on manual polysomnography scoring across nights for each study.**

| Sleep restriction study | | | | | |
|---|---|---|---|---|---|
| **Night** | **W** | **N1** | **N2** | **N3** | **REM** |
| Extension night 1 | 9.73 | 3.14 | 33.64 | 33.55 | 19.94 |
| Extension night 2 | 7.43 | 2.55 | 33.88 | 34.38 | 21.77 |
| Restriction night 1 | 9.36 | 2.46 | 23.92 | 53.65 | 10.61 |
| Restriction night 2 | 3.76 | 1.48 | 19.47 | 62.02 | 13.27 |
| **SWSD study [a]** | | | | | |
| **Night** | **W** | **N1** | **N2** | **N3** | **REM** |
| Baseline night | 8.81 | 4.83 | 42.27 | 22.20 | 21.89 |
| SWSD night | 8.86 | 6.04 | 54.04 | 8.44 | 22.61 |
| Recovery night | 7.50 | 4.62 | 42.03 | 24.04 | 21.80 |

[a] Statistical comparisons across nights have been reported in prior work using subsamples of this dataset; see (Lima Santos et al., 2025; McMakin et al., 2016; Palatine et al., 2024).

**Supplemental Table 2. Sleep restriction study – Confusion matrix for Extension Nights.**

*Supplemental Table 2A: Extension night 1* [a]

| | | Reference - Manual scoring | | | | |
|---|---|---|---|---|---|---|
| | | Wake | N1 | N2 | N3 | REM |
| **POPS** | Wake | 4776 (97.1%) | 680 (42.7%) | 642 (3.8%) | 140 (0.8%) | 455 (4.5%) |
| | N1 | 61 (1.2%) | 500 (31.4%) | 894 (5.2%) | 81 (0.5%) | 313 (3.1%) |
| | N2 | 38 (0.8%) | 160 (10.1%) | 13889 (81.5%) | 5144 (30.3%) | 520 (5.1%) |
| | N3 | 16 (0.3%) | 3 (0.2%) | 330 (1.9%) | 11486 (67.6%) | 7 (0.1%) |
| | REM | 28 (0.6%) | 249 (15.6%) | 1290 (7.6%) | 130 (0.8%) | 8819 (87.2%) |
| | | Reference - Manual scoring | | | | |
| | | Wake | N1 | N2 | N3 | REM |
| **YASA** | Wake | 4717 (95.9%) | 660 (41.5%) | 942 (5.5%) | 478 (2.8%) | 366 (3.6%) |
| | N1 | 72 (1.5%) | 400 (25.1%) | 349 (2.0%) | 29 (0.2%) | 222 (2.2%) |
| | N2 | 79 (1.6%) | 358 (22.5%) | 14607 (85.7%) | 5410 (31.9%) | 1350 (13.3%) |
| | N3 | 10 (0.2%) | 1 (0.1%) | 299 (1.8%) | 11001 (64.8%) | 5 (0.0%) |
| | REM | 41 (0.8%) | 173 (10.9%) | 848 (5.0%) | 63 (0.4%) | 8171 (80.8%) |
| | | Reference - Manual scoring | | | | |
| | | Wake | N1 | N2 | N3 | REM |
| **Combined** | Wake | 4782 (97.2%) | 700 (44.0%) | 878 (5.2%) | 345 (2.0%) | 439 (4.3%) |
| | N1 | 42 (0.9%) | 431 (27.1%) | 508 (3.0%) | 32 (0.2%) | 163 (1.6%) |
| | N2 | 53 (1.1%) | 242 (15.2%) | 14378 (84.4%) | 5237 (30.8%) | 889 (8.8%) |
| | N3 | 11 (0.2%) | 0 (0.0%) | 265 (1.6%) | 11265 (66.3%) | 5 (0.0%) |
| | REM | 31 (0.6%) | 219 (13.8%) | 1016 (6.0%) | 102 (0.6%) | 8618 (85.2%) |

*Supplemental Table 2B: Extension night 2* [a]

| | | Reference - Manual scoring | | | | |
|---|---|---|---|---|---|---|
| | | Wake | N1 | N2 | N3 | REM |
| **POPS** | Wake | 3648 (95.8%) | 505 (38.6%) | 685 (3.9%) | 161 (0.9%) | 634 (5.7%) |
| | N1 | 46 (1.2%) | 400 (30.6%) | 843 (4.9%) | 89 (0.5%) | 370 (3.3%) |
| | N2 | 64 (1.7%) | 108 (8.3%) | 14135 (81.4%) | 5481 (31.1%) | 557 (5.0%) |
| | N3 | 18 (0.5%) | 2 (0.2%) | 256 (1.5%) | 11813 (67.1%) | 13 (0.1%) |
| | REM | 30 (0.8%) | 292 (22.3%) | 1452 (8.4%) | 65 (0.4%) | 9591 (85.9%) |
| | | Reference - Manual scoring | | | | |
| | | Wake | N1 | N2 | N3 | REM |
| **YASA** | Wake | 3625 (95.2%) | 535 (40.9%) | 803 (4.6%) | 656 (3.7%) | 394 (3.5%) |
| | N1 | 47 (1.2%) | 308 (23.6%) | 357 (2.1%) | 26 (0.1%) | 330 (3.0%) |
| | N2 | 82 (2.2%) | 239 (18.3%) | 14965 (86.1%) | 5452 (31.0%) | 1534 (13.7%) |
| | N3 | 6 (0.2%) | 4 (0.3%) | 259 (1.5%) | 11429 (64.9%) | 21 (0.2%) |
| | REM | 46 (1.2%) | 221 (16.9%) | 987 (5.7%) | 46 (0.3%) | 8886 (79.6%) |
| | | Reference - Manual scoring | | | | |
| | | Wake | N1 | N2 | N3 | REM |
| **Combined** | Wake | 3663 (96.2%) | 550 (42.1%) | 789 (4.5%) | 450 (2.6%) | 535 (4.8%) |
| | N1 | 35 (0.9%) | 339 (25.9%) | 476 (2.7%) | 48 (0.3%) | 207 (1.9%) |
| | N2 | 69 (1.8%) | 151 (11.6%) | 14701 (84.6%) | 5493 (31.2%) | 974 (8.7%) |
| | N3 | 9 (0.2%) | 4 (0.3%) | 220 (1.3%) | 11571 (65.7%) | 14 (0.1%) |
| | REM | 30 (0.8%) | 263 (20.1%) | 1185 (6.8%) | 47 (0.3%) | 9435 (84.5%) |

Abbreviations: POPS – POPulation level Sleep Stager; YASA – Yet Another Spindle Algorithm.

[a] Cell values represent *n* (%) of 30-s epochs assigned to each combination of automated and manual sleep stages, pooled across all participants for the corresponding study night. Rows indicate

automated classifications, and columns indicate reference manual classifications. Percentages are calculated within manual sleep-stage categories (columns).

### Supplemental Table 3. Sleep restriction study – Confusion matrix for Restriction nights.

***Supplemental Table 3A: Restriction night 1 [a]***

| | | Reference - Manual scoring | | | | |
|---|---|---|---|---|---|---|
| | | Wake | N1 | N2 | N3 | REM |
| **POPS** | Wake | 1861 (95.5%) | 288 (56.5%) | 339 (6.8%) | 146 (1.3%) | 101 (4.6%) |
| | N1 | 49 (2.5%) | 140 (27.5%) | 460 (9.3%) | 131 (1.2%) | 75 (3.4%) |
| | N2 | 23 (1.2%) | 44 (8.6%) | 3589 (72.3%) | 3324 (29.8%) | 113 (5.1%) |
| | N3 | 9 (0.5%) | 4 (0.8%) | 29 (0.6%) | 7416 (66.6%) | 2 (0.1%) |
| | REM | 6 (0.3%) | 34 (6.7%) | 547 (11.0%) | 126 (1.1%) | 1911 (86.8%) |
| | | Reference - Manual scoring | | | | |
| | | Wake | N1 | N2 | N3 | REM |
| **YASA** | Wake | 1798 (92.3%) | 304 (59.6%) | 332 (6.7%) | 460 (4.1%) | 52 (2.4%) |
| | N1 | 14 (0.7%) | 61 (12.0%) | 70 (1.4%) | 19 (0.2%) | 46 (2.1%) |
| | N2 | 105 (5.4%) | 123 (24.1%) | 4335 (87.3%) | 3299 (29.6%) | 398 (18.1%) |
| | N3 | 17 (0.9%) | 5 (1.0%) | 39 (0.8%) | 7352 (66.0%) | 4 (0.2%) |
| | REM | 14 (0.7%) | 17 (3.3%) | 188 (3.8%) | 13 (0.1%) | 1702 (77.3%) |
| | | Reference - Manual scoring | | | | |
| | | Wake | N1 | N2 | N3 | REM |
| **Combined** | Wake | 1846 (94.8%) | 312 (61.2%) | 317 (6.4%) | 351 (3.1%) | 63 (2.9%) |
| | N1 | 15 (0.8%) | 78 (15.3%) | 172 (3.5%) | 60 (0.5%) | 30 (1.4%) |
| | N2 | 61 (3.1%) | 86 (16.9%) | 4152 (83.6%) | 3319 (29.8%) | 235 (10.7%) |
| | N3 | 14 (0.7%) | 5 (1.0%) | 28 (0.6%) | 7348 (65.9%) | 4 (0.2%) |
| | REM | 12 (0.6%) | 29 (5.7%) | 295 (5.9%) | 65 (0.6%) | 1870 (84.9%) |

***Supplemental Table 3B: Restriction night 2 [a]***

| | | Reference - Manual scoring | | | | |
|---|---|---|---|---|---|---|
| | | Wake | N1 | N2 | N3 | REM |
| **POPS** | Wake | 725 (91.8%) | 163 (53.3%) | 267 (6.6%) | 248 (1.9%) | 87 (3.2%) |
| | N1 | 19 (2.4%) | 106 (34.6%) | 453 (11.2%) | 242 (1.9%) | 115 (4.2%) |
| | N2 | 21 (2.7%) | 9 (2.9%) | 2491 (61.7%) | 3173 (24.6%) | 239 (8.7%) |
| | N3 | 21 (2.7%) | 8 (2.6%) | 77 (1.9%) | 9076 (70.3%) | 33 (1.2%) |
| | REM | 4 (0.5%) | 20 (6.5%) | 750 (18.6%) | 174 (1.3%) | 2285 (82.8%) |
| | | Reference - Manual scoring | | | | |
| | | Wake | N1 | N2 | N3 | REM |
| **YASA** | Wake | 736 (93.2%) | 221 (72.2%) | 419 (10.4%) | 489 (3.8%) | 89 (3.2%) |
| | N1 | 5 (0.6%) | 26 (8.5%) | 102 (2.5%) | 96 (0.7%) | 57 (2.1%) |
| | N2 | 22 (2.8%) | 38 (12.4%) | 3161 (78.3%) | 3451 (26.7%) | 763 (27.7%) |

| | | Wake | N1 | N2 | N3 | REM |
|---|---|---|---|---|---|---|
| | N3 | 26 (3.3%) | 8 (2.6%) | 87 (2.2%) | 8818 (68.3%) | 36 (1.3%) |
| | REM | 1 (0.1%) | 13 (4.2%) | 269 (6.7%) | 59 (0.5%) | 1814 (65.7%) |
| | | Reference - Manual scoring | | | | |
| | | Wake | N1 | N2 | N3 | REM |
| **Combined** | Wake | 735 (93.0%) | 208 (68.0%) | 352 (8.7%) | 427 (3.3%) | 84 (3.0%) |
| | N1 | 5 (0.6%) | 45 (14.7%) | 179 (4.4%) | 114 (0.9%) | 55 (2.0%) |
| | N2 | 22 (2.8%) | 25 (8.2%) | 2997 (74.2%) | 3266 (25.3%) | 450 (16.3%) |
| | N3 | 25 (3.2%) | 8 (2.6%) | 81 (2.0%) | 8999 (69.7%) | 36 (1.3%) |
| | REM | 3 (0.4%) | 20 (6.5%) | 429 (10.6%) | 107 (0.8%) | 2134 (77.3%) |

Abbreviations: POPS – POPulation level Sleep Stager; YASA – Yet Another Spindle Algorithm.

[a] Cell values represent *n* (%) of 30-s epochs assigned to each combination of automated and manual sleep stages, pooled across all participants for the corresponding study night. Rows indicate automated classifications, and columns indicate reference manual classifications. Percentages are calculated within manual sleep-stage categories (columns).

**Supplemental Table 4. Linear mixed-effects models comparing Cohen's Kappa across nights in the Sleep restriction study.**

| Contrast | Algorithm | Estimate | P [a] | FDR-corrected P [a] |
|---|---|---|---|---|
| Extension night 1 - Extension night 2 | POPS | 0.01 | 0.958 | 0.994 |
| | YASA | 0.01 | 0.994 | 0.994 |
| | Combined | 0.01 | 0.986 | 0.994 |
| Extension night 1 - Restriction night 1 | POPS | 0.11 | **<0.001** | **<0.001** |
| | YASA | 0.08 | **0.005** | **0.008** |
| | Combined | 0.09 | **0.001** | **0.002** |
| Extension night 1 - Restriction night 2 | POPS | 0.16 | **<0.001** | **<0.001** |
| | YASA | 0.16 | **<0.001** | **<0.001** |
| | Combined | 0.15 | **<0.001** | **<0.001** |
| Extension night 2 - Restriction night 1 | POPS | 0.10 | **<0.001** | **<0.001** |
| | YASA | 0.07 | **0.011** | **0.015** |
| | Combined | 0.08 | **0.002** | **0.004** |
| Extension night 2 - Restriction night 2 | POPS | 0.15 | **<0.001** | **<0.001** |
| | YASA | 0.15 | **<0.001** | **<0.001** |
| | Combined | 0.14 | **<0.001** | **<0.001** |
| Restriction night 1 - Restriction night 2 | POPS | 0.05 | 0.113 | 0.136 |
| | YASA | 0.08 | **0.004** | **0.007** |
| | Combined | 0.06 | *0.051* | *0.064* |

Abbreviations: POPS – POPulation level Sleep Stager; YASA – Yet Another Spindle Algorithm.

[a] P-values ≤ 0.05 are reported in bold characters. P-values between 0.05 and 0.10 are reported in italics.

**Supplemental Table 5. Linear mixed-effects models comparing F1 scores across nights in the Sleep restriction study.**

| **Sleep stage** | **Contrast** | **Algorithm** | **Estimate** | **P [a]** | **FDR-corrected P [a]** |
|---|---|---|---|---|---|
| Wake | Extension night 1 – Extension night 2 | POPS | 0.04 | 0.584 | 0.805 |
| | | YASA | 0.03 | 0.748 | 0.816 |
| | | Combined | 0.03 | 0.690 | 0.816 |
| | Extension night 1 – Restriction night 1 | POPS | 0.05 | 0.297 | 0.446 |
| | | YASA | 0.07 | 0.127 | 0.229 |
| | | Combined | 0.06 | 0.171 | 0.280 |
| | Extension night 1 – Restriction night 2 | POPS | 0.16 | **<0.001** | **<0.001** |
| | | YASA | 0.23 | **<0.001** | **<0.001** |
| | | Combined | 0.21 | **<0.001** | **<0.001** |
| | Extension night 2 – Restriction night 1 | POPS | 0.01 | 0.961 | 0.961 |
| | | YASA | 0.04 | 0.626 | 0.805 |
| | | Combined | 0.03 | 0.771 | 0.816 |
| | Extension night 2 – Restriction night 2 | POPS | 0.12 | **<0.001** | **0.001** |
| | | YASA | 0.20 | **<0.001** | **<0.001** |
| | | Combined | 0.18 | **<0.001** | **<0.001** |
| | Restriction night 1 – Restriction night 2 | POPS | 0.11 | **0.003** | **0.005** |
| | | YASA | 0.16 | **<0.001** | **<0.001** |
| | | Combined | 0.15 | **<0.001** | **<0.001** |
| N1 | Extension night 1 – Extension night 2 | POPS | -0.01 | 0.979 | >0.999 |
| | | YASA | 0.01 | 0.958 | >0.999 |
| | | Combined | -0.01 | 0.988 | >0.999 |
| | Extension night 1 – Restriction night 1 | POPS | 0.09 | **0.018** | **0.027** |
| | | YASA | 0.12 | **<0.001** | **0.001** |
| | | Combined | 0.13 | **<0.001** | **0.001** |
| | Extension night 1 – Restriction night 2 | POPS | 0.09 | **0.014** | **0.023** |
| | | YASA | 0.17 | **<0.001** | **<0.001** |
| | | Combined | 0.17 | **<0.001** | **<0.001** |
| | Extension night 2 – Restriction night 1 | POPS | 0.10 | **0.005** | **0.010** |
| | | YASA | 0.11 | **0.002** | **0.005** |
| | | Combined | 0.13 | **<0.001** | **<0.001** |
| | | POPS | 0.10 | **0.004** | **0.008** |
| | | YASA | 0.16 | **<0.001** | **<0.001** |

| | | | | | |
|---|---|---|---|---|---|
| | Extension night 2 – Restriction night 2 | Combined | 0.18 | **<0.001** | **<0.001** |
| | Restriction night 1 – Restriction night 2 | POPS | <0.01 | >0.999 | >0.999 |
| | | YASA | 0.05 | 0.290 | 0.401 |
| | | Combined | 0.04 | 0.513 | 0.660 |
| N2 | Extension night 1 – Extension night 2 | POPS | <0.01 | >0.999 | >0.999 |
| | | YASA | -0.01 | 0.994 | >0.999 |
| | | Combined | <0.01 | >0.999 | >0.999 |
| | Extension night 1 – Restriction night 1 | POPS | 0.17 | **<0.001** | **<0.001** |
| | | YASA | 0.11 | **<0.001** | **<0.001** |
| | | Combined | 0.13 | **<0.001** | **<0.001** |
| | Extension night 1 – Restriction night 2 | POPS | 0.27 | **<0.001** | **<0.001** |
| | | YASA | 0.22 | **<0.001** | **<0.001** |
| | | Combined | 0.23 | **<0.001** | **<0.001** |
| | Extension night 2 – Restriction night 1 | POPS | 0.17 | **<0.001** | **<0.001** |
| | | YASA | 0.12 | **<0.001** | **<0.001** |
| | | Combined | 0.13 | **<0.001** | **<0.001** |
| | Extension night 2 – Restriction night 2 | POPS | 0.27 | **<0.001** | **<0.001** |
| | | YASA | 0.23 | **<0.001** | **<0.001** |
| | | Combined | 0.23 | **<0.001** | **<0.001** |
| | Restriction night 1 – Restriction night 2 | POPS | 0.10 | **0.001** | **0.001** |
| | | YASA | 0.11 | **<0.001** | **<0.001** |
| | | Combined | 0.10 | **0.001** | **0.002** |
| N3 | Extension night 1 – Extension night 2 | POPS | 0.01 | 0.865 | >0.999 |
| | | YASA | 0.01 | 0.944 | >0.999 |
| | | Combined | 0.01 | 0.796 | >0.999 |
| | Extension night 1 – Restriction night 1 | POPS | 0.01 | 0.783 | >0.999 |
| | | YASA | <0.01 | >0.999 | >0.999 |
| | | Combined | 0.01 | 0.849 | >0.999 |
| | Extension night 1 – Restriction night 2 | POPS | -0.01 | 0.931 | >0.999 |
| | | YASA | -0.02 | 0.721 | >0.999 |
| | | Combined | -0.01 | 0.868 | >0.999 |
| | Extension night 2 – Restriction night 1 | POPS | <0.01 | 0.998 | >0.999 |
| | | YASA | -0.01 | 0.915 | >0.999 |
| | | Combined | <0.01 | >0.999 | >0.999 |
| | | POPS | -0.02 | 0.515 | >0.999 |
| | | YASA | -0.02 | 0.377 | >0.999 |

| | | | | | |
|---|---|---|---|---|---|
| | Extension night 2 – Restriction night 2 | Combined | -0.02 | 0.335 | >0.999 |
| | Restriction night 1 – Restriction night 2 | POPS | -0.02 | 0.416 | >0.999 |
| | | YASA | -0.01 | 0.773 | >0.999 |
| | | Combined | -0.02 | 0.393 | >0.999 |
| REM | Extension night 1 – Extension night 2 | POPS | 0.01 | 0.995 | >0.999 |
| | | YASA | 0.01 | 0.997 | >0.999 |
| | | Combined | <0.01 | 0.999 | >0.999 |
| | Extension night 1 – Restriction night 1 | POPS | 0.11 | **0.002** | **0.017** |
| | | YASA | 0.07 | 0.206 | 0.337 |
| | | Combined | 0.08 | *0.084* | 0.169 |
| | Extension night 1 – Restriction night 2 | POPS | 0.12 | **0.002** | **0.017** |
| | | YASA | 0.13 | **0.003** | **0.017** |
| | | Combined | 0.09 | **0.037** | 0.095 |
| | Extension night 2 – Restriction night 1 | POPS | 0.11 | **0.005** | **0.017** |
| | | YASA | 0.06 | 0.292 | 0.438 |
| | | Combined | 0.08 | 0.111 | 0.200 |
| | Extension night 2 – Restriction night 2 | POPS | 0.11 | **0.004** | **0.017** |
| | | YASA | 0.12 | **0.006** | **0.017** |
| | | Combined | 0.09 | 0.051 | 0.114 |
| | Restriction night 1 – Restriction night 2 | POPS | <0.01 | >0.999 | >0.999 |
| | | YASA | 0.06 | 0.386 | 0.534 |
| | | Combined | 0.01 | 0.986 | >0.999 |

Abbreviations: POPS – POPulation level Sleep Stager; YASA – Yet Another Spindle Algorithm.

[a] P-values ≤ 0.05 are reported in bold characters. P-values between 0.05 and 0.10 are reported in italics.

### Supplemental Table 6. Confusion matrix for SWSD study.

***Supplemental Table 6A: Baseline night*** [a]

| | | Reference - Manual scoring | | | | |
|---|---|---|---|---|---|---|
| | | Wake | N1 | N2 | N3 | REM |
| **POPS** | Wake | 1370 (89.7%) | 381 (45.3%) | 264 (3.6%) | 19 (0.5%) | 447 (11.8%) |
| | N1 | 35 (2.3%) | 144 (17.1%) | 237 (3.2%) | 3 (0.1%) | 180 (4.7%) |
| | N2 | 74 (4.8%) | 194 (23.1%) | 6135 (83.2%) | 531 (13.8%) | 146 (3.8%) |
| | N3 | 16 (1.0%) | 0 (0.0%) | 311 (4.2%) | 3297 (85.6%) | 0 (0.0%) |
| | REM | 33 (2.2%) | 122 (14.5%) | 423 (5.7%) | 0 (0.0%) | 3023 (79.6%) |

| | | Reference - Manual scoring | | | | |
|---|---|---|---|---|---|---|
| | | Wake | N1 | N2 | N3 | REM |
| **YASA** | Wake | 1315 (85.7%) | 260 (30.9%) | 128 (1.7%) | 30 (0.8%) | 79 (2.1%) |
| | N1 | 50 (3.3%) | 172 (20.5%) | 113 (1.5%) | 2 (0.1%) | 132 (3.5%) |
| | N2 | 107 (7.0%) | 279 (33.2%) | 6368 (86.4%) | 453 (11.8%) | 416 (11.0%) |
| | N3 | 15 (1.0%) | 0 (0.0%) | 484 (6.6%) | 3365 (87.4%) | 0 (0.0%) |
| | REM | 47 (3.1%) | 130 (15.5%) | 277 (3.8%) | 0 (0.0%) | 3169 (83.5%) |

| | | Reference - Manual scoring | | | | |
|---|---|---|---|---|---|---|
| | | Wake | N1 | N2 | N3 | REM |
| **Combined** | Wake | 1361 (88.7%) | 336 (40.0%) | 191 (2.6%) | 25 (0.6%) | 268 (7.1%) |
| | N1 | 33 (2.2%) | 153 (18.2%) | 113 (1.5%) | 1 (0.0%) | 86 (2.3%) |
| | N2 | 93 (6.1%) | 229 (27.2%) | 6385 (86.6%) | 472 (12.3%) | 304 (8.0%) |
| | N3 | 14 (0.9%) | 0 (0.0%) | 350 (4.7%) | 3352 (87.1%) | 0 (0.0%) |
| | REM | 33 (2.2%) | 123 (14.6%) | 331 (4.5%) | 0 (0.0%) | 3138 (82.7%) |

***Supplemental Table 6B: SWSD night*** [a]

| | | Reference - Manual scoring | | | | |
|---|---|---|---|---|---|---|
| | | Wake | N1 | N2 | N3 | REM |
| **POPS** | Wake | 1456 (93.9%) | 379 (35.9%) | 469 (5.0%) | 93 (6.4%) | 485 (12.4%) |
| | N1 | 26 (1.7%) | 190 (18.0%) | 117 (1.2%) | 0 (0.0%) | 124 (3.2%) |
| | N2 | 53 (3.4%) | 356 (33.7%) | 7984 (85.3%) | 583 (40.2%) | 99 (2.5%) |
| | N3 | 6 (0.4%) | 9 (0.9%) | 441 (4.7%) | 775 (53.4%) | 0 (0.0%) |
| | REM | 10 (0.6%) | 121 (11.5%) | 349 (3.7%) | 0 (0.0%) | 3197 (81.9%) |

| | | Reference - Manual scoring | | | | |
|---|---|---|---|---|---|---|
| | | Wake | N1 | N2 | N3 | REM |
| **YASA** | Wake | 1356 (86.9%) | 273 (25.9%) | 367 (3.9%) | 88 (6.1%) | 156 (4.0%) |
| | N1 | 75 (4.8%) | 239 (22.7%) | 113 (1.2%) | 0 (0.0%) | 83 (2.1%) |
| | N2 | 65 (4.2%) | 382 (36.2%) | 7296 (77.9%) | 347 (23.9%) | 112 (2.9%) |
| | N3 | 28 (1.8%) | 13 (1.2%) | 1178 (12.6%) | 1016 (70.0%) | 2 (0.1%) |
| | REM | 36 (2.3%) | 148 (14.0%) | 406 (4.3%) | 0 (0.0%) | 3552 (91.0%) |

| | | Reference - Manual scoring | | | | |
|---|---|---|---|---|---|---|
| | | Wake | N1 | N2 | N3 | REM |
| **Combined** | Wake | 1446 (92.7%) | 356 (33.7%) | 446 (4.8%) | 104 (7.2%) | 273 (7.0%) |
| | N1 | 19 (1.2%) | 182 (17.3%) | 86 (0.9%) | 0 (0.0%) | 51 (1.3%) |
| | N2 | 54 (3.5%) | 370 (35.1%) | 7691 (82.2%) | 425 (29.3%) | 106 (2.7%) |
| | N3 | 17 (1.1%) | 11 (1.0%) | 753 (8.0%) | 922 (63.5%) | 2 (0.1%) |
| | REM | 24 (1.5%) | 136 (12.9%) | 384 (4.1%) | 0 (0.0%) | 3473 (88.9%) |

***Supplemental Table 6C: Recovery night***

| | | Reference - Manual scoring | | | | |
|---|---|---|---|---|---|---|
| | | Wake | N1 | N2 | N3 | REM |
| **POPS** | Wake | 1204 (92.0%) | 368 (45.7%) | 292 (4.0%) | 33 (0.8%) | 654 (17.4%) |
| | N1 | 21 (1.6%) | 158 (19.6%) | 215 (3.0%) | 1 (0.0%) | 268 (7.1%) |
| | N2 | 49 (3.7%) | 127 (15.8%) | 6037 (83.0%) | 417 (10.0%) | 110 (2.9%) |
| | N3 | 15 (1.1%) | 1 (0.1%) | 235 (3.2%) | 3708 (89.2%) | 1 (0.0%) |
| | REM | 19 (1.5%) | 151 (18.8%) | 495 (6.8%) | 0 (0.0%) | 2735 (72.6%) |

| | | Reference - Manual scoring | | | | |
|---|---|---|---|---|---|---|
| | | Wake | N1 | N2 | N3 | REM |
| **YASA** | Wake | 1127 (85.8%) | 223 (27.7%) | 137 (1.9%) | 77 (1.9%) | 171 (4.5%) |
| | N1 | 39 (3.0%) | 197 (24.5%) | 108 (1.5%) | 0 (0.0%) | 148 (3.9%) |
| | N2 | 67 (5.1%) | 231 (28.7%) | 6299 (86.6%) | 314 (7.5%) | 453 (12.0%) |
| | N3 | 17 (1.3%) | 2 (0.2%) | 395 (5.4%) | 3768 (90.6%) | 2 (0.1%) |
| | REM | 63 (4.8%) | 152 (18.9%) | 335 (4.6%) | 0 (0.0%) | 2994 (79.5%) |
| | | Reference - Manual scoring | | | | |
| | | Wake | N1 | N2 | N3 | REM |
| **Combined** | Wake | 1173 (89.3%) | 328 (40.7%) | 217 (3.0%) | 43 (1.0%) | 385 (10.2%) |
| | N1 | 24 (1.8%) | 132 (16.4%) | 102 (1.4%) | 0 (0.0%) | 113 (3.0%) |
| | N2 | 61 (4.6%) | 181 (22.5%) | 6290 (86.5%) | 366 (8.8%) | 313 (8.3%) |
| | N3 | 16 (1.2%) | 1 (0.1%) | 285 (3.9%) | 3750 (90.2%) | 1 (0.0%) |
| | REM | 39 (3.0%) | 163 (20.2%) | 380 (5.2%) | 0 (0.0%) | 2956 (78.5%) |

Abbreviations: POPS – POPulation level Sleep Stager; YASA – Yet Another Spindle Algorithm; SWSD - Slow-Wave Sleep Deprivation.

[a] Cell values represent n (%) of 30-s epochs assigned to each combination of automated and manual sleep stages, pooled across all participants for the corresponding study night. Rows indicate automated classifications, and columns indicate reference manual classifications. Percentages are calculated within manual sleep-stage categories (columns).

**Supplemental Table 7. Linear mixed-effects models comparing Cohen's Kappa across nights in the SWSD study.**

| Contrast | Algorithm | Estimate | P [a] | FDR-corrected P [a] |
|---|---|---|---|---|
| Baseline night - SWSD night | POPS | 0.06 | **0.013** | **0.022** |
| | YASA | 0.09 | **<0.001** | **0.002** |
| | Combined | 0.08 | **0.002** | **0.005** |
| Baseline night - Recovery night | POPS | <0.01 | 0.987 | 0.996 |
| | YASA | <0.01 | 0.972 | 0.996 |
| | Combined | <0.01 | 0.996 | 0.996 |
| SWSD night - Recovery night | POPS | -0.06 | **0.018** | **0.027** |
| | YASA | -0.10 | **<0.001** | **0.002** |
| | Combined | -0.08 | **0.002** | **0.005** |

Abbreviations: POPS – POPulation level Sleep Stager; YASA – Yet Another Spindle Algorithm; SWSD - Slow-Wave Sleep Deprivation.

[a] P-values ≤ 0.05 are reported in bold characters.

**Supplemental Table 8. Linear mixed-effects models comparing F1 scores across nights in the SWSD study.**

| Sleep stage | Contrast | Algorithm | Estimate | P [a] | FDR-corrected P [a] |
|---|---|---|---|---|---|
| Wake | Baseline night - SWSD night | POPS | 0.04 | 0.679 | 0.918 |
| | | YASA | 0.08 | 0.117 | 0.570 |
| | | Combined | 0.05 | 0.479 | 0.862 |
| | Baseline night - Recovery night | POPS | 0.07 | 0.253 | 0.570 |
| | | YASA | 0.07 | 0.239 | 0.570 |
| | | Combined | 0.07 | 0.223 | 0.570 |
| | SWSD night - Recovery | POPS | 0.03 | 0.723 | 0.918 |
| | | YASA | -0.02 | 0.918 | 0.918 |
| | | Combined | 0.02 | 0.862 | 0.918 |
| N1 | Baseline night - SWSD night | POPS | -0.06 | 0.195 | 0.540 |
| | | YASA | -0.06 | 0.205 | 0.540 |
| | | Combined | -0.01 | 0.944 | 0.996 |
| | Baseline night - Recovery night | POPS | <0.01 | 0.996 | 0.996 |
| | | YASA | -0.02 | 0.829 | 0.996 |
| | | Combined | 0.05 | 0.392 | 0.705 |
| | SWSD night - Recovery | POPS | 0.06 | 0.227 | 0.540 |
| | | YASA | 0.04 | 0.487 | 0.731 |
| | | Combined | 0.06 | 0.240 | 0.540 |
| N2 | Baseline night - SWSD night | POPS | -0.02 | 0.184 | 0.753 |
| | | YASA | 0.02 | 0.310 | 0.753 |
| | | Combined | <0.01 | 0.932 | 0.932 |
| | Baseline night - Recovery night | POPS | -0.01 | 0.458 | 0.753 |
| | | YASA | -0.01 | 0.502 | 0.753 |
| | | Combined | -0.01 | 0.626 | 0.805 |
| | SWSD night - Recovery | POPS | 0.01 | 0.822 | 0.925 |
| | | YASA | -0.03 | **0.034** | 0.309 |
| | | Combined | -0.01 | 0.413 | 0.753 |
| N3 | Baseline night - SWSD night | POPS | 0.33 | **<0.001** | **<0.001** |
| | | YASA | 0.34 | **<0.001** | **<0.001** |
| | | Combined | 0.32 | **<0.001** | **<0.001** |
| | Baseline night - Recovery night | POPS | -0.04 | 0.417 | 0.455 |
| | | YASA | -0.04 | 0.400 | 0.455 |
| | | Combined | -0.03 | 0.455 | 0.455 |
| | SWSD night - Recovery | POPS | -0.37 | **<0.001** | **<0.001** |
| | | YASA | -0.37 | **<0.001** | **<0.001** |
| | | Combined | -0.35 | **<0.001** | **<0.001** |

| | | | | | |
|---|---|---|---|---|---|
| REM | Baseline night - SWSD night | POPS | -0.03 | 0.515 | 0.515 |
| | | YASA | -0.03 | 0.217 | 0.325 |
| | | Combined | -0.04 | 0.329 | 0.388 |
| | Baseline night - Recovery night | POPS | 0.05 | 0.139 | 0.271 |
| | | YASA | 0.04 | 0.150 | 0.271 |
| | | Combined | 0.04 | 0.345 | 0.388 |
| | SWSD night - Recovery | POPS | 0.08 | **0.012** | *0.052* |
| | | YASA | 0.07 | **0.003** | **0.024** |
| | | Combined | 0.07 | **0.019** | *0.057* |

Abbreviations: POPS – POPulation level Sleep Stager; YASA – Yet Another Spindle Algorithm; SWSD - Slow-Wave Sleep Deprivation.

[a] P-values ≤ 0.05 are reported in bold characters.

**Supplemental Table 9. Sensitivity and specificity across study nights, algorithms, and sleep stages.**

**Sleep restriction study**

| Night | Algorithm | Sensitivity | | | | | Specificity | | | | |
|---|---|---|---|---|---|---|---|---|---|---|---|
| | | Wake | N1 | N2 | N3 | REM | Wake | N1 | N2 | N3 | REM |
| Extension night 1 | POPS | 0.97 | 0.31 | 0.81 | 0.68 | 0.87 | 0.96 | 0.97 | 0.83 | 0.99 | 0.96 |
| | YASA | 0.96 | 0.25 | 0.86 | 0.65 | 0.81 | 0.95 | 0.99 | 0.79 | 0.99 | 0.97 |
| | Combined | 0.97 | 0.27 | 0.84 | 0.66 | 0.85 | 0.95 | 0.98 | 0.81 | 0.99 | 0.97 |
| Extension night 2 | POPS | 0.96 | 0.31 | 0.81 | 0.67 | 0.86 | 0.96 | 0.97 | 0.82 | 0.99 | 0.95 |
| | YASA | 0.95 | 0.24 | 0.86 | 0.65 | 0.80 | 0.95 | 0.98 | 0.78 | 0.99 | 0.97 |
| | Combined | 0.96 | 0.26 | 0.85 | 0.66 | 0.85 | 0.95 | 0.98 | 0.80 | 0.99 | 0.96 |
| Restriction night 1 | POPS | 0.96 | 0.27 | 0.72 | 0.67 | 0.87 | 0.95 | 0.96 | 0.78 | 1.00 | 0.96 |
| | YASA | 0.92 | 0.12 | 0.87 | 0.66 | 0.77 | 0.94 | 0.99 | 0.75 | 0.99 | 0.99 |
| | Combined | 0.95 | 0.15 | 0.84 | 0.66 | 0.85 | 0.94 | 0.99 | 0.77 | 0.99 | 0.98 |
| Restriction night 2 | POPS | 0.92 | 0.35 | 0.62 | 0.70 | 0.83 | 0.96 | 0.96 | 0.79 | 0.98 | 0.95 |
| | YASA | 0.93 | 0.08 | 0.78 | 0.68 | 0.66 | 0.94 | 0.99 | 0.75 | 0.98 | 0.98 |
| | Combined | 0.93 | 0.15 | 0.74 | 0.70 | 0.77 | 0.95 | 0.98 | 0.78 | 0.98 | 0.97 |

**SWSD study**

| Night | Algorithm | Sensitivity | | | | | Specificity | | | | |
|---|---|---|---|---|---|---|---|---|---|---|---|
| | | Wake | N1 | N2 | N3 | REM | Wake | N1 | N2 | N3 | REM |
| Baseline night | POPS | 0.90 | 0.17 | 0.83 | 0.86 | 0.80 | 0.93 | 0.97 | 0.91 | 0.98 | 0.96 |
| | YASA | 0.86 | 0.20 | 0.86 | 0.87 | 0.83 | 0.97 | 0.98 | 0.87 | 0.96 | 0.97 |
| | Combined | 0.89 | 0.18 | 0.87 | 0.87 | 0.83 | 0.95 | 0.99 | 0.89 | 0.97 | 0.96 |
| SWSD night | POPS | 0.94 | 0.18 | 0.85 | 0.53 | 0.82 | 0.91 | 0.98 | 0.86 | 0.97 | 0.96 |
| | YASA | 0.87 | 0.23 | 0.78 | 0.70 | 0.91 | 0.94 | 0.98 | 0.89 | 0.92 | 0.96 |
| | Combined | 0.93 | 0.17 | 0.82 | 0.64 | 0.89 | 0.93 | 0.99 | 0.88 | 0.95 | 0.96 |
| Recovery night | POPS | 0.92 | 0.20 | 0.83 | 0.89 | 0.73 | 0.92 | 0.97 | 0.93 | 0.98 | 0.95 |
| | YASA | 0.86 | 0.24 | 0.87 | 0.91 | 0.79 | 0.96 | 0.98 | 0.89 | 0.97 | 0.96 |
| | Combined | 0.89 | 0.16 | 0.86 | 0.90 | 0.78 | 0.94 | 0.99 | 0.91 | 0.98 | 0.96 |

Abbreviations: POPS – POPulation level Sleep Stager; YASA – Yet Another Spindle Algorithm; SWSD - Slow-Wave Sleep Deprivation.

**Supplemental Table 10. Comparison of automated–manual sleep-stage duration correlations across nights and algorithms in the Sleep Restriction study.**

| Sleep stage | Algorithm | Comparison (Night 1 vs Night 2) | r (Night 1) | r (Night 2) | Δr | Steiger z | P [a] | FDR-corrected P [a] |
|---|---|---|---|---|---|---|---|---|
| N1 | POPs | Extension 1 vs Extension 2 | 0.498 | 0.466 | 0.032 | 0.223 | 0.824 | 0.872 |
| | | Extension 1 vs Restriction 1 | 0.498 | 0.284 | 0.214 | 1.344 | 0.179 | 0.488 |
| | | Extension 1 vs Restriction 2 | 0.498 | 0.233 | 0.265 | 1.440 | 0.150 | 0.488 |
| | | Extension 2 vs Restriction 1 | 0.466 | 0.284 | 0.182 | 1.003 | 0.316 | 0.583 |
| | | Extension 2 vs Restriction 2 | 0.466 | 0.233 | 0.232 | 1.343 | 0.179 | 0.488 |
| | | Restriction 1 vs Restriction 2 | 0.284 | 0.233 | 0.050 | 0.279 | 0.780 | 0.867 |
| | YASA | Extension 1 vs Extension 2 | 0.682 | 0.420 | 0.263 | 2.129 | **0.033** | 0.291 |
| | | Extension 1 vs Restriction 1 | 0.682 | 0.379 | 0.303 | 2.213 | **0.027** | 0.291 |
| | | Extension 1 vs Restriction 2 | 0.682 | 0.154 | 0.528 | 3.001 | **0.003** | *0.097* |
| | | Extension 2 vs Restriction 1 | 0.420 | 0.379 | 0.041 | 0.235 | 0.815 | 0.872 |
| | | Extension 2 vs Restriction 2 | 0.420 | 0.154 | 0.266 | 1.389 | 0.165 | 0.488 |
| | | Restriction 1 vs Restriction 2 | 0.379 | 0.154 | 0.225 | 1.214 | 0.225 | 0.488 |
| | Combined | Extension 1 vs Extension 2 | 0.660 | 0.557 | 0.103 | 0.840 | 0.401 | 0.671 |
| | | Extension 1 vs Restriction 1 | 0.660 | 0.352 | 0.308 | 2.197 | **0.028** | 0.291 |
| | | Extension 1 vs Restriction 2 | 0.660 | 0.308 | 0.352 | 2.146 | **0.032** | 0.291 |
| | | Extension 2 vs Restriction 1 | 0.557 | 0.352 | 0.205 | 1.209 | 0.227 | 0.488 |
| | | Extension 2 vs Restriction 2 | 0.557 | 0.308 | 0.249 | 1.500 | 0.134 | 0.488 |
| | | Restriction 1 vs Restriction 2 | 0.352 | 0.308 | 0.044 | 0.265 | 0.791 | 0.867 |
| N2 | POPs | Extension 1 vs Extension 2 | 0.384 | 0.409 | -0.025 | -0.149 | 0.881 | 0.907 |
| | | Extension 1 vs Restriction 1 | 0.384 | 0.453 | -0.069 | -0.408 | 0.684 | 0.860 |
| | | Extension 1 vs Restriction 2 | 0.384 | 0.293 | 0.091 | 0.487 | 0.627 | 0.806 |
| | | Extension 2 vs Restriction 1 | 0.409 | 0.453 | -0.044 | -0.284 | 0.776 | 0.867 |
| | | Extension 2 vs Restriction 2 | 0.409 | 0.293 | 0.116 | 0.687 | 0.492 | 0.699 |

| | | | | | | | | |
|---|---|---|---|---|---|---|---|---|
| | | Restriction 1 vs Restriction 2 | 0.453 | 0.293 | 0.160 | 0.978 | 0.328 | 0.590 |
| | YASA | Extension 1 vs Extension 2 | 0.398 | 0.578 | -0.180 | -1.229 | 0.219 | 0.488 |
| | | Extension 1 vs Restriction 1 | 0.398 | 0.573 | -0.175 | -1.074 | 0.283 | 0.574 |
| | | Extension 1 vs Restriction 2 | 0.398 | 0.418 | -0.020 | -0.113 | 0.910 | 0.923 |
| | | Extension 2 vs Restriction 1 | 0.578 | 0.573 | 0.005 | 0.038 | 0.969 | 0.969 |
| | | Extension 2 vs Restriction 2 | 0.578 | 0.418 | 0.160 | 1.009 | 0.313 | 0.583 |
| | | Restriction 1 vs Restriction 2 | 0.573 | 0.418 | 0.155 | 1.036 | 0.300 | 0.583 |
| | Combined | Extension 1 vs Extension 2 | 0.432 | 0.547 | -0.115 | -0.758 | 0.448 | 0.673 |
| | | Extension 1 vs Restriction 1 | 0.432 | 0.622 | -0.190 | -1.221 | 0.222 | 0.488 |
| | | Extension 1 vs Restriction 2 | 0.432 | 0.475 | -0.043 | -0.260 | 0.795 | 0.867 |
| | | Extension 2 vs Restriction 1 | 0.547 | 0.622 | -0.075 | -0.577 | 0.564 | 0.781 |
| | | Extension 2 vs Restriction 2 | 0.547 | 0.475 | 0.072 | 0.489 | 0.625 | 0.806 |
| | | Restriction 1 vs Restriction 2 | 0.622 | 0.475 | 0.147 | 1.064 | 0.287 | 0.574 |
| N3 | POPs | Extension 1 vs Extension 2 | 0.487 | 0.383 | 0.104 | 0.806 | 0.420 | 0.673 |
| | | Extension 1 vs Restriction 1 | 0.487 | 0.687 | -0.200 | -1.411 | 0.158 | 0.488 |
| | | Extension 1 vs Restriction 2 | 0.487 | 0.590 | -0.103 | -0.758 | 0.448 | 0.673 |
| | | Extension 2 vs Restriction 1 | 0.383 | 0.687 | -0.304 | -2.146 | **0.032** | 0.291 |
| | | Extension 2 vs Restriction 2 | 0.383 | 0.590 | -0.207 | -1.446 | 0.148 | 0.488 |
| | | Restriction 1 vs Restriction 2 | 0.687 | 0.590 | 0.097 | 0.803 | 0.422 | 0.673 |
| | YASA | Extension 1 vs Extension 2 | 0.545 | 0.379 | 0.165 | 1.344 | 0.179 | 0.488 |
| | | Extension 1 vs Restriction 1 | 0.545 | 0.643 | -0.099 | -0.758 | 0.449 | 0.673 |
| | | Extension 1 vs Restriction 2 | 0.545 | 0.594 | -0.049 | -0.377 | 0.706 | 0.862 |
| | | Extension 2 vs Restriction 1 | 0.379 | 0.643 | -0.264 | -1.910 | *0.056* | 0.337 |
| | | Extension 2 vs Restriction 2 | 0.379 | 0.594 | -0.215 | -1.525 | 0.127 | 0.488 |
| | | Restriction 1 vs Restriction 2 | 0.643 | 0.594 | 0.049 | 0.395 | 0.693 | 0.860 |
| | Combined | Extension 1 vs Extension 2 | 0.549 | 0.396 | 0.154 | 1.263 | 0.207 | 0.488 |

| | | | | | | | | |
|---|---|---|---|---|---|---|---|---|
| | | Extension 1 vs Restriction 1 | 0.549 | 0.662 | -0.112 | -0.861 | 0.389 | 0.667 |
| | | Extension 1 vs Restriction 2 | 0.549 | 0.595 | -0.045 | -0.349 | 0.727 | 0.867 |
| | | Extension 2 vs Restriction 1 | 0.396 | 0.662 | -0.266 | -1.923 | *0.055* | 0.337 |
| | | Extension 2 vs Restriction 2 | 0.396 | 0.595 | -0.199 | -1.433 | 0.152 | 0.488 |
| | | Restriction 1 vs Restriction 2 | 0.662 | 0.595 | 0.067 | 0.541 | 0.589 | 0.800 |
| R | POPs | Extension 1 vs Extension 2 | 0.720 | 0.554 | 0.166 | 1.468 | 0.142 | 0.488 |
| | | Extension 1 vs Restriction 1 | 0.720 | 0.639 | 0.081 | 0.738 | 0.461 | 0.677 |
| | | Extension 1 vs Restriction 2 | 0.720 | 0.578 | 0.142 | 1.199 | 0.230 | 0.488 |
| | | Extension 2 vs Restriction 1 | 0.554 | 0.639 | -0.085 | -0.683 | 0.495 | 0.699 |
| | | Extension 2 vs Restriction 2 | 0.554 | 0.578 | -0.024 | -0.185 | 0.853 | 0.890 |
| | | Restriction 1 vs Restriction 2 | 0.639 | 0.578 | 0.061 | 0.501 | 0.617 | 0.806 |
| | YASA | Extension 1 vs Extension 2 | 0.616 | 0.464 | 0.152 | 1.236 | 0.216 | 0.488 |
| | | Extension 1 vs Restriction 1 | 0.616 | 0.827 | -0.211 | -2.093 | **0.036** | 0.291 |
| | | Extension 1 vs Restriction 2 | 0.616 | 0.658 | -0.042 | -0.329 | 0.742 | 0.867 |
| | | Extension 2 vs Restriction 1 | 0.464 | 0.827 | -0.363 | -3.357 | **0.001** | *0.057* |
| | | Extension 2 vs Restriction 2 | 0.464 | 0.658 | -0.194 | -1.433 | 0.152 | 0.488 |
| | | Restriction 1 vs Restriction 2 | 0.827 | 0.658 | 0.169 | 1.998 | **0.046** | 0.329 |
| | Combined | Extension 1 vs Extension 2 | 0.683 | 0.533 | 0.150 | 1.315 | 0.188 | 0.488 |
| | | Extension 1 vs Restriction 1 | 0.683 | 0.804 | -0.121 | -1.306 | 0.192 | 0.488 |
| | | Extension 1 vs Restriction 2 | 0.683 | 0.648 | 0.035 | 0.298 | 0.766 | 0.867 |
| | | Extension 2 vs Restriction 1 | 0.533 | 0.804 | -0.272 | -2.583 | **0.010** | 0.235 |
| | | Extension 2 vs Restriction 2 | 0.533 | 0.648 | -0.115 | -0.928 | 0.353 | 0.621 |
| | | Restriction 1 vs Restriction 2 | 0.804 | 0.648 | 0.157 | 1.733 | *0.083* | 0.460 |

Abbreviations: POPS – POPulation level Sleep Stager; YASA – Yet Another Spindle Algorithm.

[a] P-values ≤ 0.05 are reported in bold characters. P-values between 0.05 and 0.10 are reported in italics.

**Supplemental Table 11. Comparison of automated–manual sleep-stage duration correlations across nights and algorithms in the SWSD study.**

| Sleep stage | Algorithm | Comparison (Night 1 vs Night 2) | r (Night 1) | r (Night 2) | Δr | Steiger z | P [a] | FDR-corrected P [a] |
|---|---|---|---|---|---|---|---|---|
| | | Baseline vs SWSD | -0.123 | 0.255 | -0.378 | -1.522 | 0.128 | 0.261 |
| | POPs | Baseline vs Recovery | -0.123 | 0.152 | -0.274 | -1.226 | 0.220 | 0.330 |
| | | SWSD vs Recovery | 0.255 | 0.152 | 0.103 | 0.387 | 0.699 | 0.762 |
| | | Baseline vs SWSD | 0.154 | 0.140 | 0.014 | 0.055 | 0.956 | 0.956 |
| N1 | YASA | Baseline vs Recovery | 0.154 | 0.481 | -0.327 | -1.348 | 0.178 | 0.303 |
| | | SWSD vs Recovery | 0.140 | 0.481 | -0.341 | -1.324 | 0.185 | 0.303 |
| | | Baseline vs SWSD | 0.190 | 0.154 | 0.036 | 0.141 | 0.888 | 0.913 |
| | Combined | Baseline vs Recovery | 0.190 | 0.414 | -0.224 | -1.068 | 0.285 | 0.381 |
| | | SWSD vs Recovery | 0.154 | 0.414 | -0.260 | -0.948 | 0.343 | 0.441 |
| | | Baseline vs SWSD | 0.851 | 0.487 | 0.364 | 2.708 | **0.007** | *0.063* |
| | POPs | Baseline vs Recovery | 0.851 | 0.699 | 0.152 | 1.837 | *0.066* | 0.185 |
| | | SWSD vs Recovery | 0.487 | 0.699 | -0.212 | -1.192 | 0.233 | 0.336 |
| | | Baseline vs SWSD | 0.879 | 0.713 | 0.167 | 1.734 | *0.083* | 0.201 |
| N2 | YASA | Baseline vs Recovery | 0.879 | 0.754 | 0.126 | 1.698 | *0.090* | 0.201 |
| | | SWSD vs Recovery | 0.713 | 0.754 | -0.041 | -0.287 | 0.774 | 0.819 |
| | | Baseline vs SWSD | 0.895 | 0.659 | 0.236 | 2.416 | **0.016** | *0.094* |
| | Combined | Baseline vs Recovery | 0.895 | 0.746 | 0.149 | 2.172 | **0.030** | 0.131 |
| | | SWSD vs Recovery | 0.659 | 0.746 | -0.087 | -0.595 | 0.552 | 0.685 |
| | | Baseline vs SWSD | 0.796 | 0.436 | 0.360 | 2.094 | **0.036** | 0.131 |
| N3 | POPs | Baseline vs Recovery | 0.796 | 0.906 | -0.111 | -1.491 | 0.136 | 0.261 |
| | | SWSD vs Recovery | 0.436 | 0.906 | -0.470 | -3.046 | **0.002** | **0.042** |

| | | | | | | | | |
|---|---|---|---|---|---|---|---|---|
| | | Baseline vs SWSD | 0.789 | 0.440 | 0.349 | 2.178 | **0.029** | 0.131 |
| | YASA | Baseline vs Recovery | 0.789 | 0.875 | -0.087 | -1.103 | 0.270 | 0.374 |
| | | SWSD vs Recovery | 0.440 | 0.875 | -0.435 | -2.696 | **0.007** | *0.063* |
| | | Baseline vs SWSD | 0.834 | 0.441 | 0.393 | 2.442 | **0.015** | *0.094* |
| | Combined | Baseline vs Recovery | 0.834 | 0.920 | -0.086 | -1.388 | 0.165 | 0.297 |
| | | SWSD vs Recovery | 0.441 | 0.920 | -0.479 | -3.187 | **0.001** | **0.042** |
| | | Baseline vs SWSD | 0.320 | 0.606 | -0.286 | -1.277 | 0.201 | 0.315 |
| | POPs | Baseline vs Recovery | 0.320 | 0.165 | 0.154 | 0.548 | 0.584 | 0.701 |
| | | SWSD vs Recovery | 0.606 | 0.165 | 0.441 | 1.726 | *0.084* | 0.201 |
| | | Baseline vs SWSD | 0.497 | 0.855 | -0.358 | -2.103 | **0.035** | 0.131 |
| R | YASA | Baseline vs Recovery | 0.497 | 0.585 | -0.088 | -0.411 | 0.681 | 0.762 |
| | | SWSD vs Recovery | 0.855 | 0.585 | 0.270 | 1.834 | *0.067* | 0.185 |
| | | Baseline vs SWSD | 0.397 | 0.787 | -0.390 | -1.918 | *0.055* | 0.180 |
| | Combined | Baseline vs Recovery | 0.397 | 0.503 | -0.106 | -0.426 | 0.670 | 0.762 |
| | | SWSD vs Recovery | 0.787 | 0.503 | 0.285 | 1.485 | 0.138 | 0.261 |

Abbreviations: POPS – POPulation level Sleep Stager; YASA – Yet Another Spindle Algorithm; SWSD - Slow-Wave Sleep Deprivation.

[a] P-values ≤ 0.05 are reported in bold characters. P-values between 0.05 and 0.10 are reported in italics.

### Supplemental figures

**Supplemental Figure 1. Correlation between automated and manual sleep-stage duration estimates across nights in the Sleep Restriction study.**

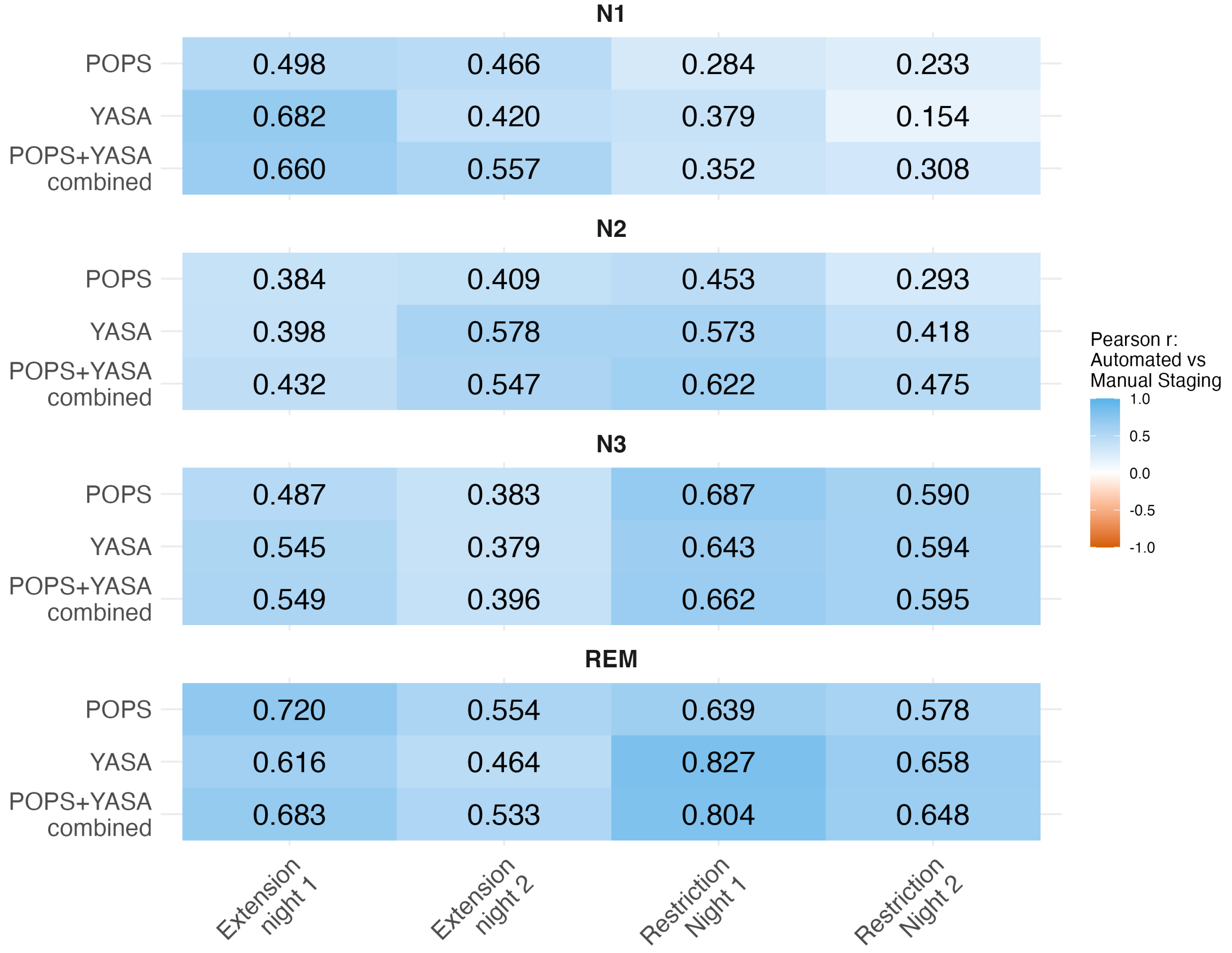


**Supplemental Figure 1 legend:** Heatmaps show Pearson correlation coefficients (*r*) between sleep-stage duration derived from manual scoring and each automated staging algorithm (POPS, YASA, and combined) for N1, N2, N3, and REM sleep in the Sleep Restriction study. Correlations were calculated separately for each night and sleep stage. Color intensity represents the magnitude and direction of the correlation, with values ranging from −1 to 1. Higher positive values indicate stronger positive associations between automated and manual stage-duration estimates.

**Supplemental Figure 2. Correlation between automated and manual sleep-stage duration estimates across nights in the SWSD study.**

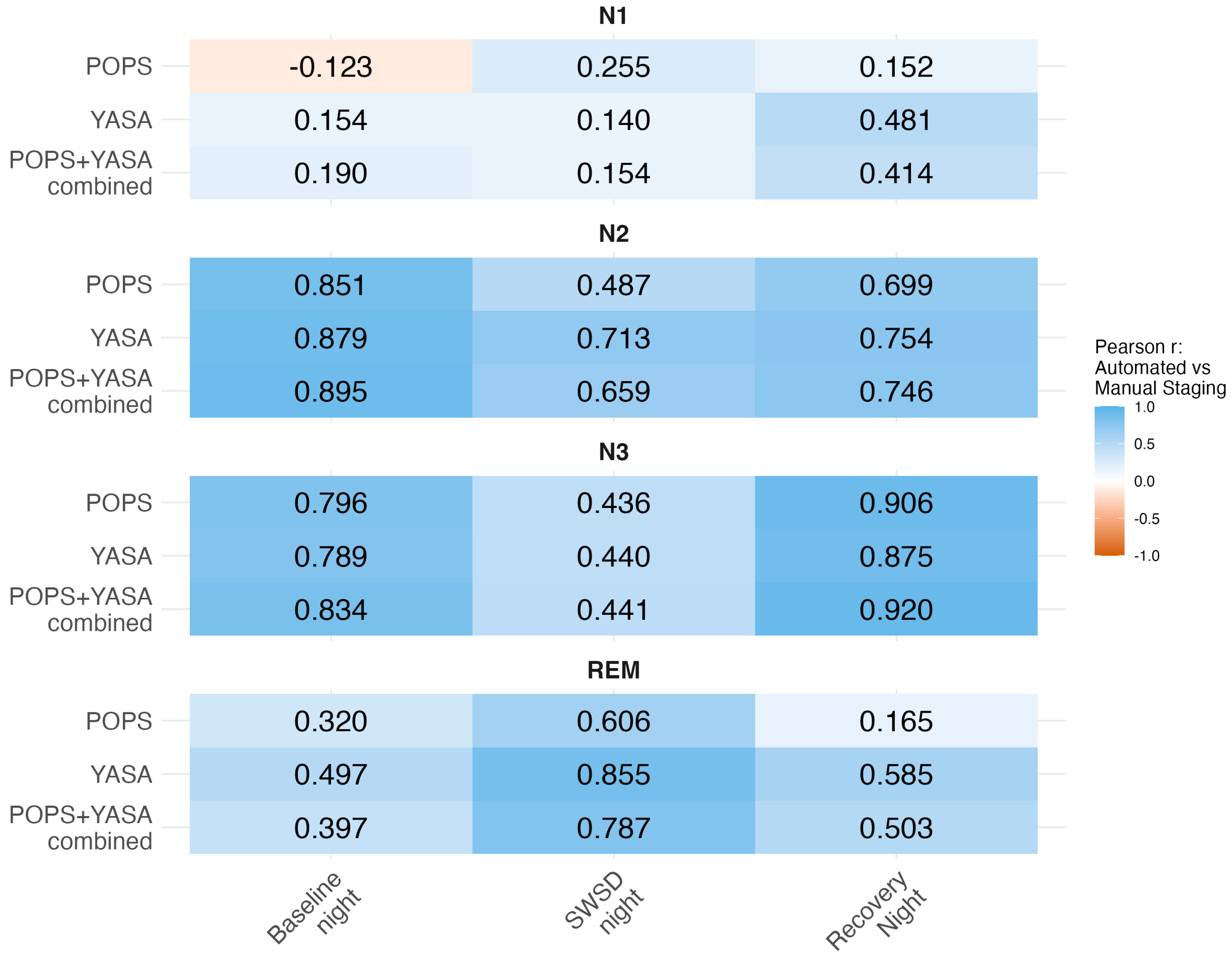


**Supplemental Figure 2 legend:** Heatmaps show Pearson correlation coefficients (*r*) between sleep-stage duration derived from manual scoring and each automated staging algorithm (POPS, YASA, and combined) for N1, N2, N3, and REM sleep in the SWSD study. Correlations were calculated separately for each night and sleep stage. Color intensity represents the magnitude and direction of the correlation, with values ranging from −1 to 1. Higher positive values indicate stronger positive associations between automated and manual stage-duration estimates.
